# A Population-Based Multi-Omics Resource for Stage I/II Melanoma Reveals Prognostic Biomarkers but Challenges Their Clinical Utility Beyond Staging

**DOI:** 10.64898/2026.11.26.26369054

**Authors:** Catherine Zhou, Thamila Kerkour, Yan-Ting Chen, Ruud W.J. Meijers, Hendrikus Jan Dubbink, Ronald van Marion, Peggy Atmodimedjo, Thierry P.P. van den Bosch, Golzar Asri Aghmuni, Luba Milena Pardo, Daphne M.S. Huigh, Lara Pozza, Erik T. Valent, Tamar Nijsten, A. Hunter Shain, Domenico Bellomo, Antien Mooyaart, Loes Hollestein

**Affiliations:** Department of Dermatology, Erasmus MC Cancer Institute, dr. Molewaterplein 40, 3015 GD Rotterdam, The Netherlands; SkylineDx B.V., Lichtenauerlaan 40, 3062 ME Rotterdam, The Netherlands; Department of Pathology and Clinical Bioinformatics, Erasmus MC Cancer Institute, Universal Medical Center Rotterdam, Rotterdam, The Netherlands; Department of Dermatology, University of California San Francisco, San Francisco, CA, USA; Helen Diller Family Comprehensive Cancer Center, University of California San Francisco, San Francisco, CA, USA

## Abstract

With the availability of adjuvant immunotherapy, improved prognostic stratification of stage I/II melanoma is needed. We investigated primary melanomas from the Dutch Early-Stage Melanoma (D-ESMEL) study. RNA and DNA sequencing was performed on 356 and 302 tumors of the discovery set, respectively. Gene expression and homozygous loss of *ARID1B* and *CDKN2A* (confirmed by p16 immunohistochemistry) were associated with metastatic risk. Findings were evaluated in the population-based validation cohort of 5,815 patients of which 306 tumors were analyzed through a nested-case control design. Although gene expression and p16 loss remained prognostic, neither improved prediction beyond clinical staging. These findings suggest that the molecular determinants of metastatic progression are largely reflected by Breslow thickness and ulceration, providing a biological explanation for the limited incremental value of molecular prognostic biomarkers in early-stage melanoma. More broadly, the D-ESMEL study establishes a framework for evaluating biomarker clinical utility through multi-omics discovery and independent population-based validation.

## Introduction

Accurate prognostication in early-stage cutaneous melanoma is critical for guiding clinical management and identifying patients at risk of progression. The American Joint Committee on Cancer (AJCC) staging system, based on Breslow thickness and ulceration, has demonstrated robust prognostic utility^1^. Nevertheless, these clinicopathological parameters alone do not fully capture the biological heterogeneity that drives variability in patient outcomes in early-stage disease, as a subset of patients still develops distant metastases despite favorable initial staging^2–4^. Approximately, 41% of melanoma deaths and 60% of distant metastases occur in patients initially diagnosed with stage I/II disease^2,4^. This underscores the need for molecular biomarkers based on the excised primary melanoma that extend beyond current staging parameters to better capture the drivers of progression in early-stage melanoma, as this concerns almost 90% of all newly diagnosed patients.

Development and validation of clinically useful prognostic models for early-stage melanoma require large, well-powered studies with sufficient metastatic events to ensure stable, generalizable predictions and to demonstrate clinical utility ^5,6^. This poses a large challenge for stage I melanoma, for multiple reasons. First, distant metastasis after a stage I melanoma is very rare (i.e. ∼1%) and require efficient study design for multi-omics analyses^7^. Second, stage I melanomas are usually diagnosed several years before the metastasis in general hospitals, which makes it difficult for researchers from academic centers to obtain archived tissue of a sufficiently large sample for prognostic studies. Third, even within stage I melanoma, variations in Breslow thickness and ulceration are prognostic. While multivariable adjustments can address this issue, application to cohorts with limited events risks statistical instability, model overfitting, and wide confidence intervals^8,9^. As a result, one of the critical concerns of recently developed gene expression profiles (GEP) for melanoma prognosis is insufficient adjustment for established prognostic variables, which may result in high-risk GEP classifications that primarily reflect known high-risk features rather than providing independent prognostic information^10^.

The Dutch Early-Stage Melanoma (D-ESMEL) study was designed to address these challenges by evaluating the value of multi-omics analyses for identifying early-stage melanoma patients at risk of distant metastasis beyond well-established clinicopathological factors. We employed a matched case-control discovery dataset to control for key prognostic variables, followed by validation in a nationwide cohort using a nested case-control design to enable robust performance assessment^11^. Using these study designs, we sought to develop and validate prognostic models for predicting distant metastasis risk in early-stage melanoma and determine whether it provides added value over current clinicopathological staging factors.

## Results

### Discovery dataset

The clinical characteristics of the discovery dataset have previously been described in detail^11^. In summary, the discovery set is a matched case-control study, including 221 primary stage I/II melanomas that led to distant metastatic disease and 221 primary stage I/I melanomas with the same Breslow thickness, ulceration status, age, sex, and follow-up time, but without distant metastasis (Figure 1A). Of all 221 case-control sets (442 samples), 101 case-control sets (202 samples) were stage I melanoma^11^. Of all 442 samples, DNA sequencing was generated for 302 stage I/II melanomas and RNA sequencing was generated for 356 stage I/II melanomas. The distribution of patient and tumor characteristics was comparable to the entire discovery dataset (Supplementary Tables 1 and 2).

**Figure 1:**
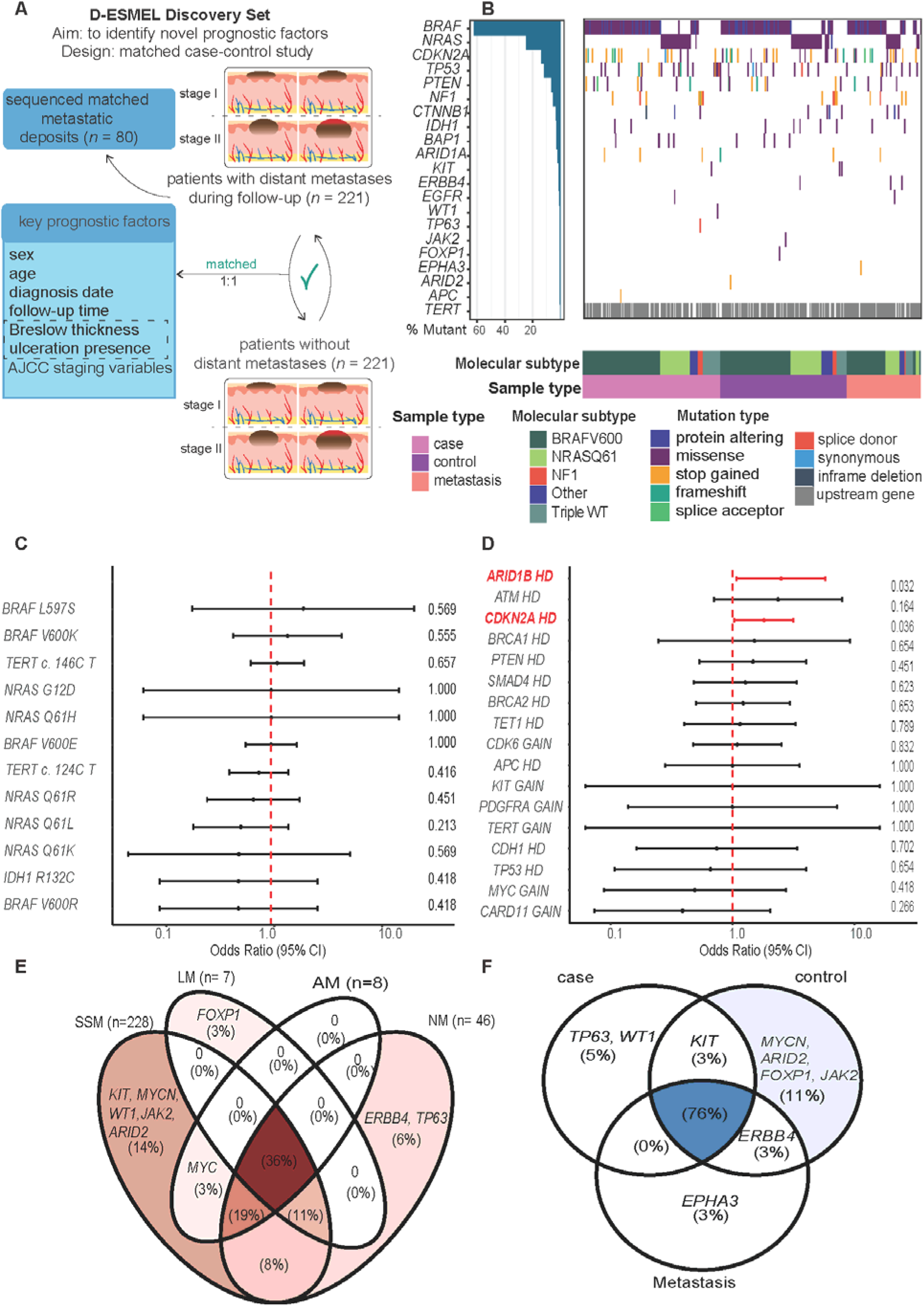
Genomic alterations in the D-ESMEL discovery dataset. **(A)** The D-ESMEL discovery set is a matched case-control study, comprising of 221 primary melanomas from patients who developed distant metastases (cases) and 221 matched controls (patients without distant metastases during the same follow-up time as the case). Distant metastatic tissue was sequenced as well for 80 cases **(B)** Distribution and frequency of somatic mutations across the analyzed samples. The left panel shows the percentage of samples harboring mutations in each gene. The central oncoplot displays the mutation profiles of individual samples. Annotation tracks indicate sample type and molecular subtype. Colors represent different mutation subtypes. **(C)** Forest plot of the regression analyses evaluating the associations between each somatic mutation and metastatic outcome. Odds ratios (ORs) and 95% confidence intervals (CIs) are shown. The red dashed vertical line indicates no association (OR = 1). **(D)** Forest plot of the regression analyses evaluating the associations between copy-number variations (CNVs) and metastatic risk. HD indicates a homozygous deletion. Odds ratios and 95% confidence intervals are shown. The red dashed vertical line indicates no association (OR = 1). **(E)** Distribution and overlap of mutated genes across histological melanoma subtypes, including (SSM) superficial spreading melanoma (SSM), lentigo maligna melanoma (LM), acral melanoma, and (NM) nodular melanoma. The diagram illustrates shared and subtype-specific mutation patterns. **(F)** Distribution and overlap of mutated genes across sample types: primary melanomas from cases and controls, and matched metastatic deposits. The diagram highlights shared and sample-type-specific mutation patterns.

### Genomic alterations associated with distant metastasis

Targeted DNA sequencing with a melanoma-specific panel designed for detecting genomic alterations of prognosis was applied (Supplementary Table 3). Targeted sequencing was considered the most feasible sequencing method because DNA was isolated from old archived FFPE material of primarily thin melanomas, for which the amount and quality of the DNA were expected to be low. Expected frequencies of molecular subtypes (BRAF mutant, RAS mutant, NF1 mutant, Triple WT) were observed, but these did not differ between primary melanomas of cases and controls (X-squared = 0.98, p-value = 0.91) (Figure1B). Point mutations were not prognostic (Figure 1C). In contrast, homozygous loss of *CDKN2A* and *ARID1B* were associated with an increased risk of distant metastasis (Figure 1D). In a combined analysis of point mutations and copy number variation, the same pattern was observed: an inactivation (i.e., a homozygous deletion, two distinct pathogenic mutations on separate alleles, or a loss of heterozygosity and a pathogenic mutation) of *CDKN2A* and *ARID1B* was associated with a twice as high risk of distant metastasis (Supplementary Tables 4 and 5).

While many mutations were shared between primary melanomas that metastasized and those that did not, some mutations were only observed in cases (*TP63, WT1*) and others only in controls (*MYCN*, *ARID2*, *FOXP1*, *JAK2*). Mutations in *EPHA3* were only observed in metastatic tissue (Figure 1E). Well-known mutations in melanoma (e.g., *TERT*, *TP53*, *BRAF*, *CDKN2A*, *ARID1B*) were shared across histological subtypes, but we also observed specific mutations per histological subtype, such as KIT, MYCN, WT1, JAK2, ARID2, which only occurred in superficial spreading melanomas, and ERBB4 and TP63 in nodular melanomas (Figure 1F).

### Clonality analyses

Of 80 patients with metastatic sequencing data, 59 also had matched primary melanoma sequencing. Clonal relatedness could not be assessed in 37% (22/59) due to limited DNA quantity or poor copy-number data quality. Clonal relatedness could be assessed in 37 patients. Within these, the majority (36/37) of the primary melanoma and the patient-matched metastasis were classified as clonally related or likely clonally related, demonstrating a high degree of genomic concordance between primary tumors and their corresponding metastases (Supplementary Table S6). Only one pair (case-control set 304) showed evidence of absence of clonal relatedness, due to presence of a *BRAF* p.Val600Lys mutation in the primary tumor that was not detected in the patient-matched metastasis.

### Transcriptomic patterns associated with distant metastasis

Consensus clustering of the gene expression patterns of 356 primary stage I/II melanomas with known genomic alterations revealed 2 gene expression clusters (Figure 2A). These clusters were associated with clinical stage, ulceration, histological subtype, and mitotic index (all P<0.0000001), but not with genomic subtype (p=0.11). Cluster 1 was associated with stage II, ulcerated, nodular melanomas with mitosis. Cluster 2 was associated with stage I melanoma, non-ulcerated, superficial spreading melanomas without mitosis. Overrepresentation analyses revealed that in cluster 1 genes involved in the EMT pathway were upregulated, while cluster 2 showed increased expression of genes involved in estrogen response, UV response, and response to hypoxia (Figure 2B). Cluster 1 was associated with poor distant-metastasis-free survival (p=0.01, Figure 2C)

**Figure 2.**
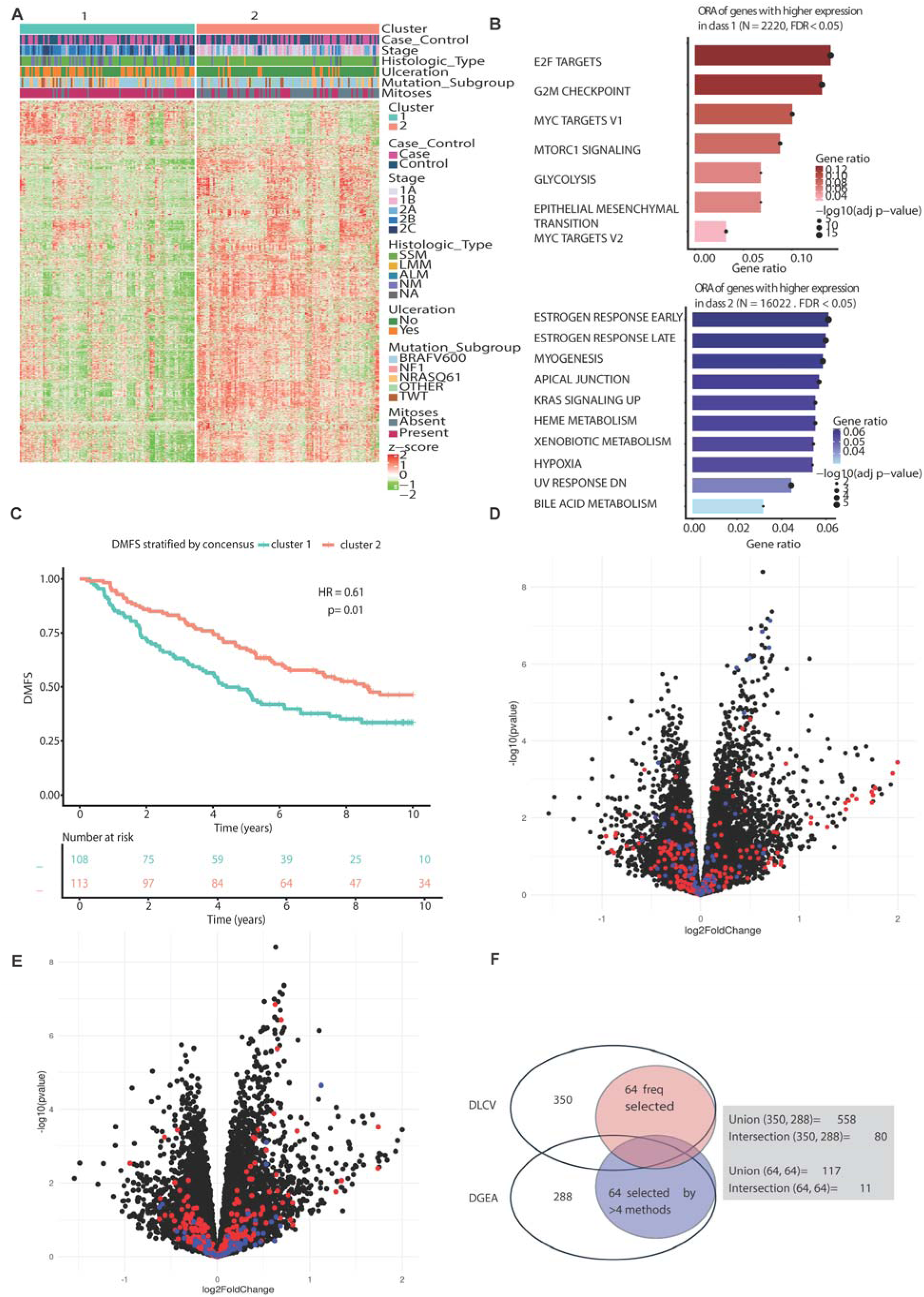
Gene expression the identified of potentially prognostic genes in the D-ESMEL discovery set. **(A)** Unsupervised consensus clustering of RNA-sequencing data from primary melanoma samples in the discovery set. The heatmap displays gene expression levels as row-wise z-scores, with red and blue indicating relatively higher and lower expression, respectively. Annotation tracks indicate case or control, AJCC stage, histological subtype, ulceration status, genomic subtype and mitosis. **(B)** Overrepresentation analysis of genes upregulated in Cluster 1 and Cluster 2. Enriched biological processes and pathways are shown for each cluster. Gene ratio represented by bar length, false discovery rate (FDR) by color intensity, and gene count by point size. **(C)** Kaplan Meier curve of distant metastasis free survival, stratified by consensus cluster. **(D)** Volcano plot showing differential gene expression identified using a double-loop cross-validation (DLCV) approach. The x-axis represents log₂ fold change, and the y-axis represents −log₁₀-transformed P values. Upregulated genes are shown in red, downregulated genes in blue. **(E)** Volcano plot showing differential gene expression analysis (DGEA). The x-axis represents log₂ fold change, and the y-axis represents −log₁₀-transformed P values. Upregulated genes are shown in red, downregulated genes in blue. **(F)** Venn diagram illustrating the overlap between the identified prognostic genes by double-loop cross-validation (DLCV), differential gene expression analysis (DGEA), and survival-based selection. Numbers indicate the number of genes in each intersection or analysis-specific subset. The overlap highlights genes consistently identified across complementary analytical approaches.

A dual approach was performed to identify prognostic gene expression profiles based on differential gene expression analysis and regression modelling. First, LASSO conditional logistic regression with double-loop cross-validation resulted in 350 genes that were most frequently (defined as > 20%) selected (Figure 1D, Supplementary Figure 3). Permutation testing with 100 randomized case-control label assignments (Supplementary Figure 4) demonstrated that models built on original data performed better than the models based on permuted data, with original data models achieving an AUC of ∼0.6 with 500 or more input genes (Extended Data Figure 1A).

Second, we applied a union-based selection approach across multiple statistical models, where the selection was based on the top 100 genes with the smallest p-values from seven differential expression models (Figure 1E, Extended Data Figure 1B). Taking the overlap into account between the different models, we identified 288 differentially expressed genes between cases and controls, of which 64 were selected by >4 different methods (Figure 1F).

Combining both approaches, we constructed a final gene set comprising 558 unique genes, including 80 overlapping entries (Figure 1F, Supplementary Table 7). Among the 64 most frequently selected genes from both methods, 11 genes overlapped.

The discovery set lacked source population data due to its case-control design. Therefore prognostic modelling based on the 558 unique genes was performed in the validation cohort, where the nested case-control design enabled weighted regression analyses resulting in absolute risk prediction.

### A homozygous loss of CDKN2A correlates with a loss of P16

To validate the genomic findings at the protein level, p16 clonal loss was scored based on immunohistochemistry in 264 samples from the discovery dataset (132 matched case-control sets). Clonal loss was defined as a discrete cluster of tumor cells lacking p16 staining and was quantified as the percentage of total tumor area with loss (Figure 3A). Each increase of 10% loss of p16 staining was associated with distant metastasis (OR 1.07, 95% CI 1.01–1.13; P = 0.019). A binary cutoff of >20% clonal loss was used to facilitate implementation in clinical practice and also showed that >20% clonal loss of p16 staining was more common in cases (81/132, 61%) than in matched controls (58/132, 44%) (OR 2.05, 95% CI 1.23–3.41; P = 0.006). Despite DNA being isolated from macrodissected tumor and RNA from whole tissue, *CDKN2A* inactivation in the tumor correlated with lower whole-tissue *CDKN2A* expression (Figure 3C). p16 protein loss in the tumor did not correlate with whole-slide *CDKN2A* expression, which is consistent with the known heterogeneity of P16 expression within tissue (Figure 3D). A homozygous loss of *CDKN2A* at the genomic level correlated with P16 loss at the protein level (p = 0.016; Figure 3E), supporting p16 immunohistochemistry as a feasible surrogate for *CDKN2A* inactivation.

**Figure 3:**
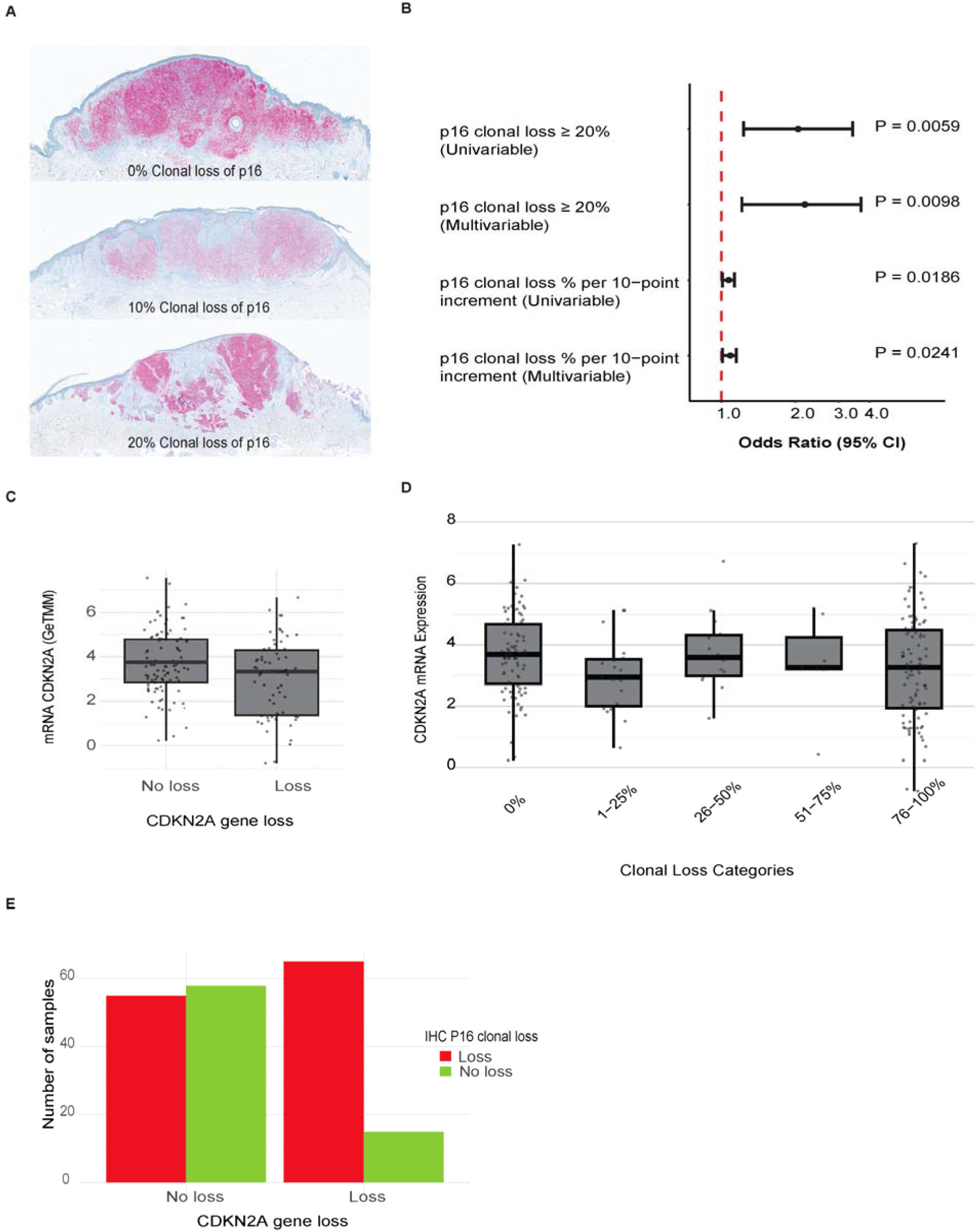
CKN2A/p16 alterations and correlation between genomic alteration and transcriptomic and protein expression in the D-ESMEL discovery set. **(A)** Representative immunohistochemical (IHC) images illustrating different levels of clonal loss of p16 expression in melanoma tissue, including 0%, 10%, and 20% clonal loss. **(B)** Forest plots of the regression analysis evaluating the association between clonal p16 loss and metastatic risk. Breslow thickness, ulceration, age and sex are controlled for in univariable and multivariable analyses by matching. Multivariable analyses are also adjusted for histological subtype, body site and mitosis **(C)** Boxplots of the association between CDKN2A DNA-level loss and CDKN2A mRNA expression. **(D)** Boxplots of the association between CDKN2A mRNA expression and p16 clonal loss assessed by IHC. **(E)** Distribution of p16 clonal loss assessed by IHC according to CDKN2A gene-loss status.

### Validation cohort

To validate our findings, a nationwide cohort of 5,851 stage I/II patients based on data from the Netherlands cancer registry was selected. A nested case-control (NCC) design was applied, including both randomly selected controls and controls matched on stage (Figure 4A)^11,12^. The first NCC dataset included 154 patients who developed distant metastasis (cases) and 154 randomly selected controls (n=154)^11^. From all 154 cases, 153 cases were also matched to a control based on AJCC substage (i.e., IA, IB, IIA, IIB, IIC) to generate a second NCC dataset^12^. The multi-omics analyses from the discovery set provided valuable insights into tumor biology, but are unlikely to be used simultaneously in clinical practice. Therefore, we validated the most feasible biomarkers for clinical practice, such as the loss of P16, as this showed to be a good surrogate marker for the most prognostic genomic finding (homozygous loss of *CDKN2A*), and developed a gene expression profile based on the 558 prognostic genes from the discovery set.

**Figure 4:**
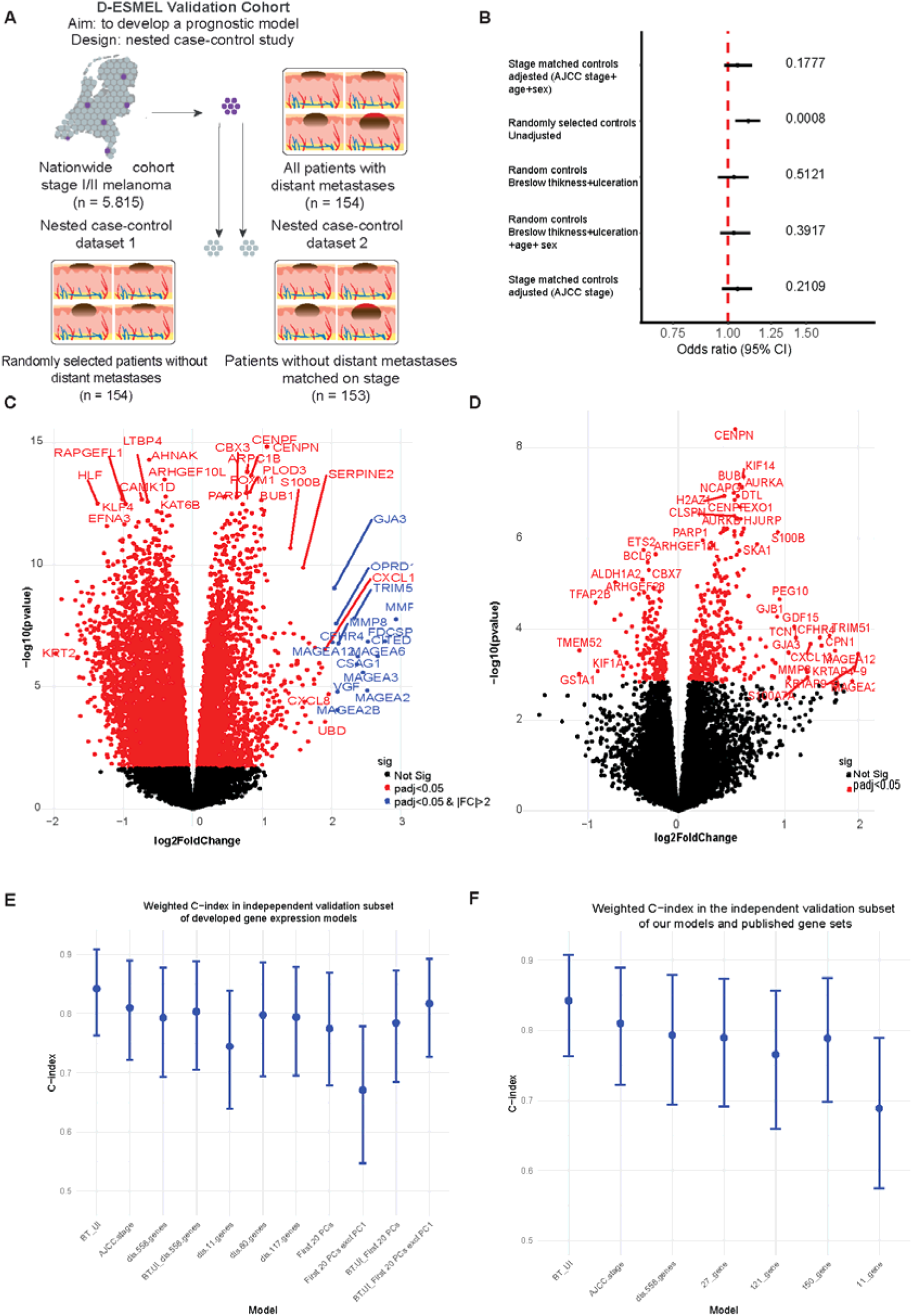
Clonal loss of p16 and gene expression in the D-ESMEL validation cohort. **(A)** The D-ESMEL validation cohort is a nested case-control study design: each case (n=154) from a population-based cohort of 5,815 stage I/II melanomas has been sampled and matched to both a randomly selected control (n=154) and a control matched on AJCC substage (n=153). This design enables weighting of the controls according to their occurrence in the source population and facilitates the development of population-based prognostic models. **(B)** Conditional logistic regression of clonal loss of p16 per 10% increment assessed by immunohistochemistry. Univariable analyses with randomly selected controls evaluate the effect of clonal loss of p16 only. Multivariable analyses are adjusted for Breslow thickness, ulceration, age and sex. Univariable analyses with stage-matched controls are controlled for AJCC substage by design. Multivariable analyses are also adjusted for age and sex. **(C)** Volcano plot of differential gene expression, unconditional analyses (limma or edgeR). Red and blue (Fold change [FC] >2) dots indicate statistically significant differentially expressed genes between cases and randomly selected controls **(D)** Volcano plot of differential gene expression, conditional analyses on matched pairs (limma or edgeR). Red dots indicate statistically significant differentially expressed genes between cases and randomly selected controls **(E)** Weighted c-index of prognostic models in the independent validation subset (1/3 of the total cohort, which was withheld during model development). The baseline prognostic models included Breslow thickness and ulceration (BT_Ul) or AJCC substage. These models were compared to models based on genes following from the discovery set (dis) and models based on principal component (PC) analyses. Bars indicate 95% CI **(F)** Weighted c-index of our prognostic models (BT_Ul, AJCC_stage, dis_558 genes) and previously published gene sets. Bars indicate 95% CI

### Validation of loss of p16 with immunohistochemistry

P16 clonal loss was evaluated in both NCC datasets (i.e., 136/154 cases, 136/154 randomly selected controls, and 136/153 stage-matched controls). NCC dataset 1 (randomly selected controls) provided insight into the prognostic value of clonal loss of p16 alone, and NCC dataset 2 (stage-matched controls) provided more statistical power in stage II^12^. Clonal loss of p16 was associated with distant metastasis (OR for ≥20% clonal loss: 2.36, 95% CI: 1.48-3.77, p=0.0003). However, clonal loss of p16 was not an independent prognostic factor after adjustment for staging variables or after matching on AJCC substage (Figure 4B, Extended Data Figure 2A). In stage II subgroups and ulcerated tumors, clonal loss of p16 also did not show independent prognostic value (Extended Data Figure 2B). As a consequence, p16 immunohistochemistry did not result in increased discriminative performance in the population-based validation cohort, as indicated by the weighted c-index, which was 0.82 for a model that included both AJCC stage and p16 immunohistochemistry (for both 20% clonal loss and per 10% loss) compared to 0.81 for AJCC stage alone.

### Model development and validation of a gene expression profile

As immunohistochemistry analyses in both NCC datasets resulted in similar outcomes, we decided to perform the gene expression analyses in NCC dataset 1 (randomly selected controls), resulting in a population-based gene expression resource of stage I/II melanomas without progression. RNA-seq was available for 306 samples (153 cases and 153 randomly selected controls). First, model development was conducted in two-thirds of the validation cohort (hereafter referred to as the model development subset) to assess generalizability and predictive accuracy of the gene set (Supplementary Table 8). The best-performing and most robust models from the model development subset were applied to the remaining one-third of the validation cohort (hereafter referred to as the independent validation subset) to evaluate their predictive performance, enabling assessment of model stability, potential overfitting, and reproducibility in an independent sample.

First, we performed differential gene expression analysis without conditioning on known prognostic variables (i.e., age, sex, Breslow thickness, and ulceration), which resulted in 8001 differentially expressed genes between primary melanomas that led to metastasis and those that did not (Figure 4C). When conditioning on these known prognostic variables, 526 genes were differentially expressed, indicating that failure to account for these known variables results in a high proportion of differentially expressed genes that are related to known clinical and histopathological variables (Figure 4D). Therefore, all subsequent analyses were conditioned on the matched variables or the paired nature of the data.

In both the model development subset and independent validation subset, models with Breslow thickness and ulceration or AJCC stage achieved high weighted C-index (>0.80, Figure 4E and 4F, Extended Data Figure 3, Supplementary Table 9). First, a model was developed based on the 558 genes identified in the discovery dataset, which achieved a weighted C-index of 0.79 (95% CI 0.69-0.88) in the independent validation subset (Figure 4E, Supplementary Table 9). Adding the 558 genes to Breslow thickness and ulceration did not improve performance (weighted AUC 0.80 [95% CI, 0.74-0.88], weighted C-index 0.83 [95% CI, 0.77-0.89]) compared to Breslow thickness and ulceration alone (Figure 4E, Extended Data Figure 3C, Supplementary Table 7). Various gene selection methods via regularization were explored, but neither adaptive LASSO nor ridge regression improved predictive accuracy. We also tested whether using subsets of genes with higher selection frequency in the discovery set (n = 11, 80, or 117) would enhance performance, but no improvement was observed (Figure 2F & 4E, Extended Data Figure 3, Supplementary Table 9).

### Association between gene expression and clinicopathological variables

To further explore the extent to which the 558 discovery genes reflected established prognostic variables, we first examined their direct association with Breslow thickness and ulceration. We observed that 75% of the genes were associated with ulceration and 79% with Breslow thickness in the discovery set. In the model development subset of the validation cohort, 96% of genes correlated with both variables. The aggregated prediction score based on the 558 genes showed a strong correlation with Breslow thickness (r = 0.74) and significantly higher values in ulcerated compared with non-ulcerated tumors (p = 1.1 × 10 ¹) (Extended Data Figure 4A and 4B).

We then conducted principal component analysis (PCA) on the 558 discovery genes to further evaluate their independent contribution. PC1 was strongly associated with clinical stage, indicating overlap with known prognostic features (Extended Data Figure 4C). Adding clinical predictors to models based on the remaining PCs (excluding PC1) did not improve predictive performance, suggesting that GEP adds limited independent signal (Supplementary Table 9, Figure 4C). PCA-derived risk scores based on the remaining components appeared largely orthogonal to the clinical model of Breslow thickness and ulceration, implying that gene expression may reflect similar underlying biology.

We conducted exploratory analyses of previously published gene sets within the model development subset ^8,13–16^. None of these sets showed a distinct differential expression pattern in volcano plots (Extended Data Figure 5A). Predictive models built from these published gene sets demonstrated discriminative performance similar to that of our 558-gene set, and comparable to models based on Breslow thickness with ulceration or on AJCC stage (Figure 4F, Extended Data Figure 5B).

### Biological pathways analyses

Genomic alterations of oncogenic signaling pathways in early-stage melanoma showed that telomere, RAS pathway, and the cell cycle pathway were frequently altered (Figure 5A). The cell cycle pathway was more frequently altered in primary melanomas of patients who developed distant metastasis and their matched metastatic tissue compared to primary melanomas of patients without metastasis (OR=1.84, 95%: 1.13-3.01) (Supplementary Table 10).

**Figure 5.**
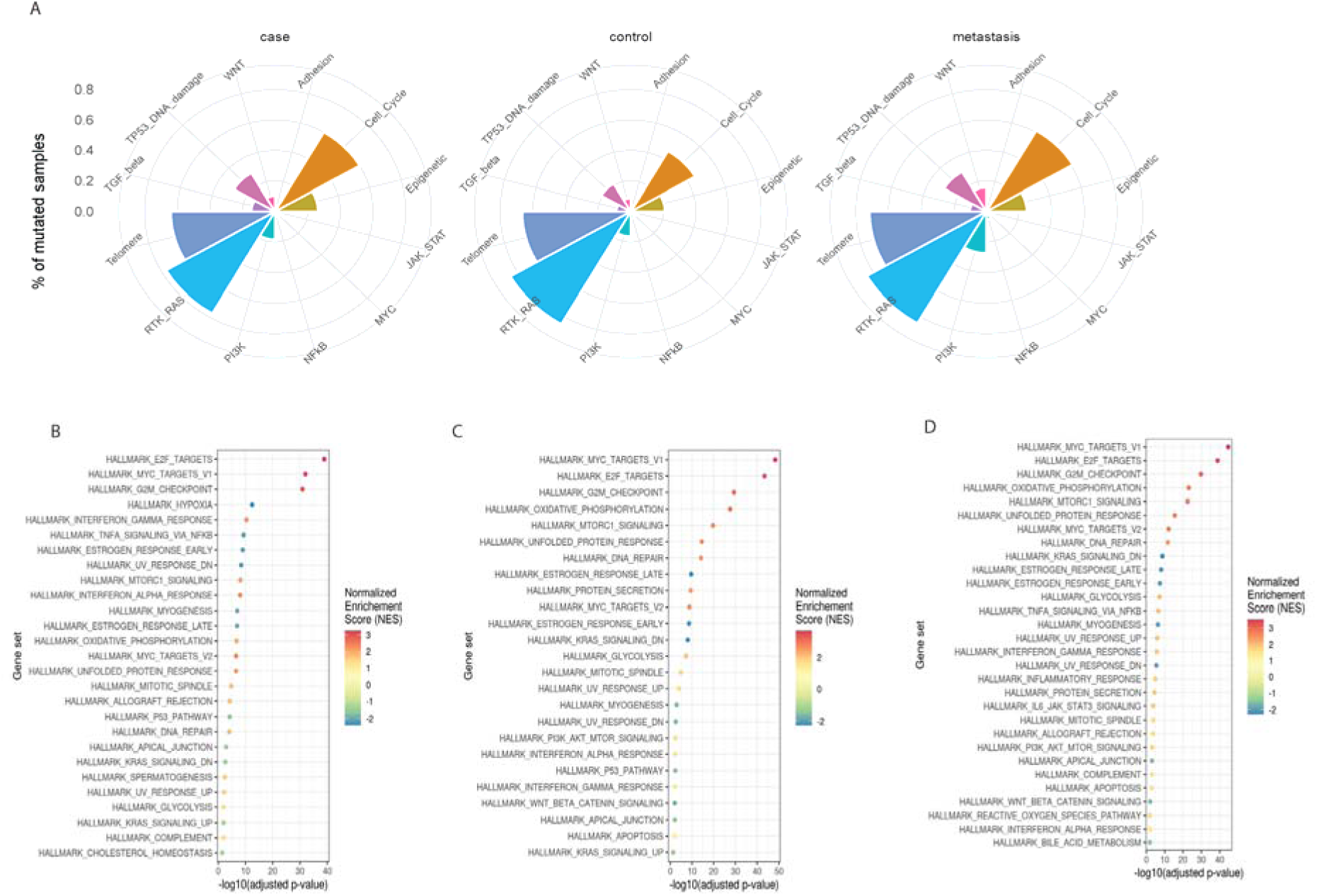
Pathway and gene set enrichment analyses in the discovery set and validation cohort of the D-ESMEL study. **(A)** Pathway analysis of DNA alterations in the D-ESMEL discovery set. Radar plots show the percentage of mutated samples across cancer-related pathways in cases, controls, and metastatic lesions. The pathways include cell cycle, apoptosis, epigenetic regulation, JAK–STAT, MYC, NF-κB, PI3K, RTK–RAS, telomere maintenance, TGF-β, and TP53/DNA damage response, Wingless/Integrated signaling pathway (WNT). **(B)** Gene set enrichment analysis (GSEA) comparing primary melanomas from cases and controls in the validation cohort, accounting for the matching variables. Gene sets are ranked by statistical significance. **(C)** GSEA comparing thin melanomas (Breslow thickness ≤1 mm) with thick melanomas (Breslow thickness >1 mm) in the validation cohort. Gene sets are ranked by statistical significance. **(D)** GSEA comparing ulcerated and non-ulcerated primary melanomas in the validation cohort. Gene sets are ranked by statistical significance.

To explore biological processes on a transcriptomic level that were associated with metastatic progression beyond individual gene effects, gene set enrichment analyses in the model development subset of the validation cohort were performed. When conditioned on the matching variables, several hallmark gene sets were significantly enriched in metastatic cases. The top enriched pathways included E2F targets, MYC targets (V1), and the G2M checkpoint, all showing strong positive enrichment with high normalized enrichment scores (NES). Additional enrichment was seen for hypoxia, estrogen response, unfolded protein response, and both interferon gamma and alpha signaling (Figure 5B).

To assess whether these signals overlap with established clinicopathological risk factors, we repeated the analyses stratified by Breslow thickness and ulceration. In thicker melanomas (>2 mm), the same proliferative programs (MYC targets, E2F targets, G2M checkpoint) were again among the top enriched gene sets, accompanied by oxidative phosphorylation, DNA repair, and mTORC1 signaling (Figure 5C). Similarly, in ulcerated melanomas, a broad set of hallmark pathways was enriched, including proliferative signaling (E2F, MYC, G2M), metabolic activity (glycolysis, oxidative phosphorylation, fatty acid metabolism), and immune responses (inflammatory response, interferon signaling) (Figure 5D).

## Discussion

The D-ESMEL study is the largest multi-omics characterization of (thin) early-stage primary melanomas that led to distant metastasis^11^. Its unique population-based design provides a roadmap for studying rare cancer outcomes and enables the development of absolute risk prediction models with potential clinical relevance ^7,17,18^. We observed that gene expression and homozygous loss of *ARID1B* and *CDKN2A* (confirmed with p16 immunohistochemistry) were associated with metastatic risk, but gene expression and loss of p16 did not outperform staging in the validation cohort. Our findings show that molecular biomarkers of early-stage melanoma largely recapitulate the prognostic information captured by Breslow thickness and ulceration, explaining why molecular signatures have struggled to outperform conventional staging.

Loss of *CDKN2A* represents a well-established event in melanoma, affecting both the p16-RB and p14-p53 pathways^19^. Loss of p16 removes a key cell-cycle checkpoint, allowing melanocytes to proliferate and driving a transition towards a more invasive, dedifferentiated phenotype. Biallelic *CDKN2A* loss represents a relatively late event in melanoma evolution and has been associated with increased *BRN2* expression, suggesting a role in phenotypic switching and melanoma progression ^20^. Previous studies also showed that a loss of p16 was associated with increased proliferation, sentinel lymph node positivity and poor survival ^21–23^. We also identified a homozygous loss of *ARID1B* as a potential predictor of metastatic progression. *ARID1B* is a component of the SWI/SNF complex and is essential for maintaining chromatin accessibility and regulating transcriptional programs^24^. Disruption of SWI/SNF components has been implicated in tumor progression across multiple cancer types^25^. However, *ARID1B* is not a known tumor suppressor gene in melanoma and is reported to be mutated in ∼10% of melanomas^26^. Possibly, a loss of chromatin remodeling capacity may facilitate transcriptional reprogramming required for invasion and metastatic colonization. Possibly, the complete chromosomal arm (6q) may have been lost, because primary melanomas with chromosomal aberrations detectable by FISH for chromosomes 6 and 11 were more likely to metastasize^27^.

Large scale sequencing studies in cutaneous melanoma including The Cancer Genome Atlas (TCGA)^28^, the Leeds melanoma cohort^8^, and the InterMEL study^29^, have expanded our knowledge on the genomic and transcriptomic landscape and defining the molecular subtypes of cutaneous melanoma. However, they either primarily included metastatic melanoma tissues, did not include stage I melanoma or the event rate among stage I was too low for adequate statistical power for prognostic modelling. In addition to these studies, several molecular signatures have been developed for melanoma prognostication, with prior evaluations reporting consistent risk stratification. We compared our gene expression profile to previously published gene sets, which were designed for predicting a high risk on relapse. CP-GEP (SkylineDx) was designed to predict a low risk on sentinel lymph node metastasis and has demonstrated high negative predictive value (95%) for sentinel lymph node positivity in a meta-analysis, supporting its potential role in SLNB decision-making^30^. A systematic review of DecisionDx-Melanoma (Castle Biosciences) showed significantly different survival outcomes across risk classes^31^. Prospective trials such as NivoMela (NCT04309409) are evaluating the utility of MelaGenix (NeraCare) in guiding adjuvant therapy. However, the clinical utility of prognostic signatures requires adequate study design and statistical power. Previous studies reported promising associations, but low event rates and wide confidence intervals limited the precision of their estimates, as illustrated by DecisionDx-Melanoma and LMC_150 ^8^. Similarly, a recently published study describing a six-gene copy number signature (including loss of *CDKN2A*) included only 37 stage I melanomas, limiting statistical power in this subgroup ^32^. Moreover, categorizing Breslow thickness in multivariable analyses may have resulted in inadequate adjustment for this key prognostic factor, potentially explaining the additional prognostic value attributed to gene expression or copy number loss ^14,32^. Alternatively, a more objective assessment of copy number loss through DNA sequencing compared to p16 loss in our study may explain this signature’s independent prognostic value. Future DNA sequencing of the D-ESMEL validation cohort will establish its added clinical utility beyond staging at the population level. Aforementioned methodological limitations may partly explain why previously published signatures appeared to provide prognostic information beyond conventional staging. These examples also highlight the importance of study design, outcome definitions, and event rates when evaluating the added value of prognostic signatures. Our study illustrates the methodological standard required before new molecular biomarkers can plausibly change clinical management in early-stage melanoma. This standard includes matched designs that control for the dominant histopathological variables, validation in population-based cohorts with sufficient events, weighted performance metrics that reflect the source population rather than the sampled case-control set, and head-to-head comparison with the established clinical model of Breslow thickness and ulceration.

Gene set enrichment analyses of our gene expression profile explained why gene expression signatures did not yield stronger predictive models that outperformed AJCC staging. Despite tight matching, many genes remained correlated with Breslow thickness and ulceration, because matching balances variables between cases and controls but does not remove their intrinsic biological links with gene expression. Gene set enrichment analyses revealed consistent enrichment of proliferative programs (E2F, MYC, G2M checkpoint), alongside hypoxia, estrogen response, unfolded protein response, and interferon signaling: pathways widely associated with tumor aggressiveness and progression^33–37^. This finding aligns with prior studies demonstrating upregulation of DNA repair and cell-cycle genes in thicker tumours^38^, and proliferative, vascular, and inflammatory profiles in ulcerated tumours^39^. The same core programs that drive thickness and ulceration, including cell-cycle disinhibition via *CDKN2A* loss, phenotype switching, and proliferative transcriptional states, are what molecular assays detect. The implication for next-generation biomarker development is that gains in prognostic performance in early-stage disease will have to come from measurements that capture biology orthogonal to tumor-cell–intrinsic aggressiveness. Potential orthogonal molecular features include tumor-immune cell interactions, such as the spatial distribution of TILs in general or more specifically the cytotoxic T-cel ^40,41^.

While our study design and use of a nationwide cancer registry facilitated long-term follow-up of a large population-based sample, the downside of this design is that old archived FFPE tumour samples resulted in a low amount and poor quality DNA, limiting the ability to perform whole exome sequencing. To account for the low amount and quality, we opted for a targeted sequencing method that is feasible with extremely low input DNA (i.e., 1 ng), which resulted in a high (70%) success rate of DNAseq of primarily thin and old samples (i.e. 50% of samples were melanomas of <1 mm and/or >10 years old).

Our findings have direct implications for the current debate on molecular biomarkers in early-stage melanoma. In a nationwide, population-based validation cohort, which is the clinical context in which prognostic tests would be deployed, neither the gene expression profile derived from our matched discovery analysis nor the previously published commercial gene sets outperformed Breslow thickness and ulceration for the prediction of distant metastasis. Similarly, p16 clonal loss did not retain prognostic value beyond staging. These findings reinforce current recommendations from the NCCN, SSO, and AAD against routine use of commercial GEP assays for prognostication or management decisions in stage I/II melanoma, and extend this conclusion to a broader molecular readout than has previously been evaluated head-to-head against staging in a weighted, population-based framework.

To conclude, the D-ESMEL study represents a unique population-based resource, combining multi-omics data with long-term clinical follow-up in the largest worldwide collection of stage I/II primary melanomas. As a consequence of our population-based study design, our findings have direct and immediate translational relevance. Our study challenges the clinical utility of prognostic biomarkers, as we showed, that they capture overlapping tumor biology of histopathological features through different measurements.

## Online methods

### Study design and patient population

This study utilized the methodological framework of the D-ESMEL study, drawing data from the Netherlands Cancer Registry (NCR) and the Dutch Nationwide Pathology Databank (PALGA), as previously described^11^. The discovery set comprised 221 matched case-control pairs of stage I/II melanoma patients matched on age, sex, Breslow thickness, and ulceration. Cases were patients who developed distant metastases; controls remained distant metastases-free within the follow-up period during which their matched case progressed. This design enabled identification of candidate genes independent of known clinicopathological factors.

The validation cohort used a nested case-control (NCC) design within a nationwide cohort of 5,815 patients diagnosed with stage I/II melanoma between 2016 and 2020, including 154 case-control sets matched on follow-up time. 153 cases were also matched to a second control based on AJCC stage^12^. The validation cohort was split into two-thirds for model development and one-third for independent validation. The NCC design permits estimation of absolute risks^7,42^.

This study was approved by the Erasmus MC Ethics Committee (MEC-2018-1738), the scientific committee of the NCR and the Privacy Committee of PALGA. The code of conduct for responsible use of human residual tissue by the Federation of Dutch Medical Scientific Societies was followed^43^.

### DNA extraction and targeted panel sequencing

The dermatopathologist (AM) selected the tumor regions for macrodissection. Tissue sections were deparaffinized and stained with hematoxylin before the microdissection of the tumor area. DNA extraction was performed in samples with a minimum tumor percentage of 30% using the Chelex method, as described previously ^44^.

A targeted DNA sequencing panel was designed to capture all potentially prognostic genomic alterations in melanoma. First, a comprehensive literature review covering studies published between 2000 and 2021, focusing on next generation sequencing studies of primary cutaneous melanomas and treatment-naïve metastatic tissues, minimize confounding effects of therapy on mutation status. We retrieved publicly available mutation data from five studies via cBioPortal^45–49^, compiling an initial list of 248 genes found to be mutated in either primary or metastatic melanoma samples (305 and 835 patients respectively). Of these genes, 27 genes overlapped with the existing diagnostic panel used at Erasmus MC for melanoma patients and were retained, as they represent established melanoma driver genes. For the remaining 221 genes, we applied additional filtering criteria to prioritize those with known roles in cancer progression and melanomagenesis. This involved mapping each gene to relevant biological pathways and evaluating potential mechanistic links to melanoma metastasis. Resources such as Cancer Hotspots, OncoKB, and NCBI databases were utilized to identify mutation hotspots and regions of interest within each gene. In total, 62 genes were included in the final targeted panel (Supplementary Table 3).

In order to prevent batch effect, samples from the same sets were processed in the same batch. All the samples with available DNA was sequenced. For NGS analysis, library preparation was performed using the Ion AmpliSeq Plus workflow, with 10 ng of input DNA per pool. In cases where less than 10 ng DNA was available, the number of PCR cycles was increased accordingly. Following library construction, template preparation was carried out using the Ion Chef system according to the manufacturer’s instructions. Sequencing was subsequently performed on the Ion GeneStudio S5 platform utilizing an Ion 540 chip.

### DNA mutations

Sequence Pilot software Version [5.2.0] (JSI medical systems GmbH, Ettenheim, Germany) was used to make a preselection of the true variants. For each amplicon, the total coverage should be >100 reads, and the strand bias coverage <0.9, the variant allele frequency (VAF) >8%. All nonsynonymous somatic points mutations, splice site alterations, insertions and deletions were included. Variants identified as common polymorphism in 1000 genomes databases in more than 1% were excluded. Variants that were present in at least 10% of the called were included. In addition, correlation between the tumor percentage and the VAF of hotspot mutation was take into account. To avoid detection of FFPE artifacts, true pathogenic/likely pathogenic variants as detected with Sequence Pilot software were validated by an molecular biologist with extensive experience in melanoma diagnostics [RM]. For the *TERT* promoter, strand bias filter was set to 0 which means variants only present in forward or reverse reads are scored as well.

We classified the samples into five molecular subtypes in melanoma, as previously reported ^50^. BRAF subtype: if mutations occurring at the hotspot position V600, NRAS subtype: if mutations was detected in the hotspot potion Q61, NF subtype: if mutation occurring on the *NF1* gene, Triple Wild Type (TWT) subtype: if no mutations on *BRAF, NRAS* or *NF1*, and finally, other if the patient had mutations on *BRAF* or *NRAS* at other locations than V600 or Q61.

### DNA copy number variation

CNVs were analyzed using a combined analyses of allelic imbalance and SNP analyses. Allelic imbalance was detected by generating the Ratio of Patient Coverage (RPC) with the Sequence Pilot software. The RPC is calculated as follows:

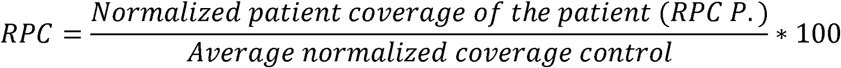

Normalization was performed using normal patient tissues from any tissue type of 15 diagnostic routine samples to correct for technical variation. The RPC value ranged from -100 (loss) to infinity (gain). An RPC-value of 0 means no CNV alterations. The median RPC of at least three amplicons per gene was used in CNV analyses. SNPs within and around each gene were used to calculate the B allele frequency and categorize them into homozygous (B-allele frequency <0.05 or >0.95), heterozygous (B-allele frequency between 0.45-0.55), and imbalanced SNP (B allele frequency between 0.05 -0.45 or 0.55-0.95). Based on the RPC value of at least 3 amplicons and imbalance of at least 2 SNPs, genes were categorized as homozygous deletion, loss of heterozygosity, or gain (Supplementary Table 11).

### DNA Pathways analysis

Gene alterations within each pathway were analysed. Major signaling pathways, which are frequently altered across cancers based on TCGA analyses, were included: RTK–RAS, PI3K, TP53/DNA damage response, cell cycle, WNT, TGF-β, and MYC signaling ^51^. In addition, we included functionally relevant pathways in melanoma: epigenetic regulation, NF-κB signaling, cell adhesion, telomere maintenance, and JAK/STAT signaling mapped by g.profiler using Gene ontology database. A pathway was considered altered if there was at least one activating oncogene or inactivating tumor suppressor gene within that pathway.

### Comparison of primary melanoma and matched metastasis

Clonal relatedness between the primary melanoma and the matched metastasis was analysed based on both copy number variations and mutations (Supplementary Table 12). Matched CNVs were prioritized, because CNVs tend to be retained during cancer progression. Thereafter, identical non-hotspot mutations and (absence of) hotspot mutations were assessed to determine clonal relatedness.

### Genomic prognostic models in the discovery set

Genes were classified according to their functional roles as either tumor suppressors or oncogenes. Tumor suppressor genes were considered inactivated if they harbored a homozygous deletion, or a loss of heterozygosity combined with a pathogenic mutation, or two distinct pathogenic mutations likely occurring on separate alleles with variant allele frequencies of approximately 40–50%. Oncogenes were classified as activated if they harbored a gain or carried a single pathogenic variant.

An unconditional logistic regression was performed with distant metastasis as dependent variable, which enabled us to take all sequenced samples into account, including those without a matching case or control. Independent variables included all matching variables (i.e. age, sex, Breslow thickness, ulceration and follow-up time) and the genomic alteration of interest. Multivariable models including all point mutations, all CNVs or (in)activation on the gene level were constructed using backward selection based on the Akaike Information Criterion (AIC) to identify alterations that predicted distant metastasis. To assess the effect of the broken matching, a sensitivity analysis was performed in a subset of complete case-control sets.

### P16 immunohistochemistry

Immunohistochemistry was performed on a Ventana Benchmark ULTRA automated staining platform using ultraview red detection kit. Following deparaffinization and heat-induced antigen retrieval, sections were incubated with anti-p16 antibody (clone E6H4, 1.23 µg ml−1; Ventana®) for 28 minutes, followed by haematoxylin II counter stain for 20 min and then a blue colouring reagent for 8 min according to the manufacturer’s instructions (Ventana® Medical Systems Inc., Arizona, USA).

### P16 Scoring and Evaluation

Clonal loss was defined as a discrete cluster of tumor cells lacking P16 staining. Clonal loss was quantified as a percentage of the total tumor area, ranging from 0% (no loss) to 100% (complete loss). Heterozygous staining was assessed as 0% clonal loss. p16 clonal loss was analyzed in parallel in the validation cohort with randomly selected controls and the validation cohort with stage-matched controls, mirroring the matched design of the discovery analyses. The primary endpoint was dichotomized p16 clonal loss (>20%, as defined in the discovery set); the paired difference in continuous clonal loss percentage was the secondary endpoint. Conditional odds ratios with 95% confidence intervals were derived from the discordant pair counts. Multivariable analyses used conditional logistic regression stratified on case-control set, with a pre-specified hierarchy: (i) unadjusted, (ii) adjusted for Breslow thickness and ulceration (the variables controlled for in the discovery-set matching), and (iii) fully adjusted with additional covariates age and sex (mirroring the complete discovery-set matching structure). Pre-specified subgroup analyses in the stage-matched NCC dataset included stage II overall (IIA/IIB/IIC), stage IIB+IIC (current adjuvant-therapy indication), stage IIC only, and ulcerated tumors; subgroups were restricted to pairs where both members qualified. A Cox proportional hazards model was applied to the NCC dataset with the stage-matched controls to evaluate prognostic models with stage and stage combined with p16 clonal loss. The proportional hazard assumption was assessed using Schoenfeld residuals. Discriminative ability was evaluated by weighted C-index, an adaptation of the C-index adjusted to the NCC design^7^.

### RNA Extraction, Library Preparation, and Sequencing

Total RNA was extracted using the RNeasy FFPE kit (Qiagen, Hilden, Germany) according to the manufacturer’s protocol. RNA concentration and purity were determined using a Nanodrop 2000 spectrophotometer (Thermo Scientific). For library preparation, 50 ng of total RNA was processed using the RNA Prep with Enrichment kit (Illumina, San Diego, CA), incorporating an exome capture approach. The concentration of the generated libraries was measured using a Qubit™ fluorometer (Thermo Fisher Scientific), libraries were pooled and sequencing was performed using the Illumina NextSeq™ system (Illumina, San Diego, CA). The system generated binary base call (BCL) files, which contained base calls for each cycle across all clusters on the flow cell, along with an interop metric file providing sequencing run statistics.

### RNAseq - Bioinformatics Processing and Quality Control

The first step in data processing involved converting BCL files to FASTQ files, followed by demultiplexing using bcl2fastq2 v2.20 (Illumina, San Diego, CA). An in-house analysis pipeline, composed of two separate software packages was then applied. The first software package generated STAR genome and Salmon transcriptome indices, while the second handled FASTQ file processing, including adapter and quality trimming, read mapping, gene expression quantification, and quality metric extraction.

The Salmon reference transcriptome was constructed from the intersection of the UCSC GoldenPath hg38 reference genome with the Gencode v38 gene model. Sequencing adapters and read segments with a Q-score below 20 were trimmed using TrimGalore. QC metrics from FastQC and GATK were aggregated, and samples identified as outliers or failing thresholds underwent further review.

Principal component analysis (PCA) (stats package v4.2.1; *plotPCA* function in DESeq2 v1.36.0) was used to assess batch effects across library preparations, sequencing runs, and pathology laboratories. When batch effects were detected, the ComBat method was applied for correction.

Read alignment was performed using STAR (RRID:SCR_004463), generating binary sequence alignment (BAM) files. Read location distribution (coding, intronic, untranslated regions (UTR), intergenic, ribosomal) and transcript coverage normalization were assessed using Picard/GATK CollectRnaSeqMetrics. Gene expression quantification was conducted using Salmon, producing counts and Transcripts-Per-Million (TPM) for each transcript. Gene-level expression values were obtained using the tximport package.

The versions of software used in Hawkweed and Foxglove (v2.04) included STAR (v2.7.10a) ^52^, Salmon (v1.9.0) ^53^, FastQC (v0.11.9, RRID:SCR_014583) ^54^, TrimGalore (v0.6.6), cutadapt (v3.4), seqtk (v1.3), GATK (4.2.0.0, RRID:SCR_001876) and samtools/htslib (v1.12) ^55^ in R version 4.4.0 ((R Project for Statistical Computing, RRID:SCR_001905).

### Differential gene expression and modelling in the discovery set

To identify a set of genes that could distinguish cases from controls, we applied two complementary analytical approaches on the discovery set: differential gene expression analysis (DGEA) and discriminative modelling.

DGEA was conducted using multiple statistical frameworks, including DESeq2 (v1.36.0, RRID:SCR_015687), edgeR (v.3.22, RRID:SCR_012802), limma (v3.62.2, RRID:SCR_010943), and direct linear regression (lm from stats v4.4.0) to assess differential expression. Each method was tested with statistical adjustment for the matched design (matching variables (all methods) and case-control pairs (excluding DESeq2, due to unstable dispersion estimates caused by the high number of case-control pairs)). ^56^ To derive a final set of differentially expressed genes for further model development, a union-based selection approach was applied. From each method, the 100 genes with the smallest *p*-values were selected. Rather than relying on a single statistical model, the final DGEA-based set was determined by taking the union of all selected genes across models.

In parallel, we used conditional logistic regression (CLR) as the basis for discriminative modelling, allowing for internal validation of the selected gene set. Feature selection and parameter estimation were performed using a penalized maximum likelihood estimation algorithm with least absolute shrinkage and selection operator (LASSO) to prevent overfitting and improve model generalization ^57,58^.To provide a robust and unbiased assessment of model performance, double-loop cross-validation (DLCV) was implemented (Supplementary Figure 3). DLCV consisted of an outer loop for model selection and an inner loop for tuning the LASSO penalty. Paired t-tests were used within each training set to rank genes by significance, and the top 5–1000 genes were used as input features to a conditional logistic regression model. The LASSO penalty parameter was optimized via 10-fold inner cross-validation. Model accuracy, as area under the curve (AUC), was evaluated by repeating this process 50 times. To determine whether the observed performance and gene selection were due to chance, case-control labels were permuted 100 times, allowing identification of genes associated with metastases selected with significantly higher frequency. (Supplementary Figure 3).

### Transcriptomic prognostic modelling in the model development subset of the validation cohort

Following identification of candidate genes in the discovery set, model development was conducted in the model development subset (i.e. two-thirds of the validation cohort) to assess generalizability and predictive accuracy of the gene set. First, we performed DGEA using DESeq2^56^ and limma^59^ to determine how many genes identified in the discovery set remained differentially expressed in the model development subset. Models were conditioned on the matching variables from the discovery set and AJCC stage.

We constructed baseline models using logistic regression with LASSO regularization: (1) clinicopathological variables (Breslow thickness and ulceration), (2) AJCC stage, (3) matching variables, (4) gene expression data, and (5) an integrated model combining clinical variables and gene expression. All genes identified in the discovery set were initially included as input features. Alternative feature selection strategies were explored, including adaptive LASSO and ridge regression. Additionally, we examined whether subsets of 558 genes with higher selection frequency across models improved predictive accuracy.

Discriminative performance was assessed using both weighted AUC and weighted C-index with corresponding 95% CI, with sampling weights applied to account for the NCC design^42^. These weights were based on the distribution of follow-up time, as controls were randomly sampled without matching on prognostic factors. Internal validation used 200 bootstrap iterations with the 0.632+ estimator^60^.

Models were trained using 200 bootstrap iterations with LASSO regularization across input sets of 50 to 1000 genes with the highest variances. Performance was assessed using weighted AUC and weighted C-index in the model development subset and the independent validation subset.

### Validation of transcriptomic prognostic models in the independent validation subset of the validation cohort

The best-performing and most robust models from the model development subset were applied to the remaining one-third of the validation cohort (i.e. the independent validation subset), which was withheld during model development. Weighted AUC and C-index with corresponding 95% CI were recalculated using the methods previously described. Sampling weights were applied based on the corresponding one-third of the nationwide source cohort.

In addition to validating our gene expression model, we conducted exploratory analyses using previously published gene sets as positive methodological controls^8,13–16^. Although these gene signatures differ in endpoints, technical platforms, and intended patient populations compared with our study, we hypothesized that they should show association with distant metastasis in our cohort, thereby serving as an internal validation of our methodology. We examined the distribution of differentially expressed genes from each signature using volcano plots. Each gene set was then used as input for logistic regression models with LASSO regularization and performance evaluated using weighted AUC and weighted C-index metrics with corresponding 95% CI in the model development subset.

### Gene set enrichment analyses

To investigate the biological pathways associated with metastatic progression and established prognostic factors, we performed gene set enrichment analysis using the MSigDB Hallmark gene sets^61^ and the DGEA on the model development subset of the validation cohort. Three complementary analyses were performed: (1) cases versus controls conditioned on matching variables, (2) melanomas with Breslow thickness >2 mm versus ≤2 mm, and (3) melanomas with versus without ulceration. Genes were ranked by moderated t-statistics from limma, and enrichment scores were calculated using the fgsea algorithm. Significance of enrichment was assessed based on normalized enrichment scores (NES) and adjusted p-values (false discovery rate <0.05).

### Statistical analyses

General descriptive statistics were used to summarize the patients’ clinical characteristics. Clinicopathological characteristics were summarized using counts and percentages for categorical variables and medians with interquartile ranges (IQR) for continuous variables. Comparisons between groups were performed using Fisher’s exact or chi-squared tests, as appropriate. Oncoplot was generated using GenVisR package version (1.42.2). Correlation between *CDKN2A* loss and p16 clonal loss within each sample was tested using Chi-square test. All statistical analyses and figure generation were performed in R (version 4.4.0). A two-sided p-value of 0.05 was considered statistically significant

## Acknowledgments

We thank all dedicated registrars of the NCR and Palga for their work on the data collection. We thank the Erasmus MC Pathology Research and Trial Service (PARTS) for sectioning the tissue material. The project was co-funded by the PPP Allowance made available by Health∼Holland (Grant Number: EMCLSH19008), Top Sector Life Sciences & Health and the Dutch Cancer Society (KWF, grant number: 2021-1-13470). Roche Diagnostics provided the reagents and technical support that enabled the immunohistochemistry analysis in this study as part of an investigator-initiated study.

## Ethics Statement

The use of left-over diagnostic tissue samples for scientific research in the Netherlands is based on the ‘no objection’ principle, as stated in the Code of Conduct for Health Research from the Committee on Regulation of Health Research. Consequently, a waiver of informed consent was granted by the Medical Ethics Review Committee of the Erasmus Medical (MEC-2018-1738). The study was also approved by the scientific and privacy committees of the NCR and Palga.

## Code Availability

Code is available on GitHub: https://github.com/erasmusmc-dermatology.

## Contributors

The project was conceptualized by AM, LH, CZ, TK, DB, HJD. Sample and clinical data collection were performed by CZ, TK, TvdB. DNA sequencing was performed by RvM, PA, TK, HJD and the resulting data was analyzed by TK, RM, AM. RNA-sequencing was performed by DH and the data was analyzed by YT-C, LaP, DB, CZ, TK. P16 immunohistochemistry was performed by TvdB and the data was analyzed by GAA, AM, CZ. Pathology revision was performed by AM. Visualization was performed by TK, CZ, YT-C, LaP, GAA. Data interpretation was performed CZ, TK, YT-C, RM, HJD, RvM, PA, GAA, LaP, EV, TN, AHS, DB, AM, LH. The work was supervised by DB, TN, AM, LH. The original draft was written by CZ, TK, LH. All authors contributed to editing and critical review of the manuscript.

## Data availability

Raw DNA and RNA sequencing data were made available through the European Genome Archive (Accession numbers EGAxxx and EGAxxx). Clinical data according to the Human Tumor Atlas Network (HTAN) phase 2 data model is available upon request at the Netherlands Cancer Registry with the following description: “HTAN dataset from data request numbers: K19.037 (D-ESMEL discovery set) and K21.277 (D-ESMEL validation cohort)”.

