## Extended data for "A Population-Based Multi-Omics Resource for Stage I/II Melanoma Reveals Prognostic Biomarkers but Challenges Their Clinical Utility Beyond Staging"

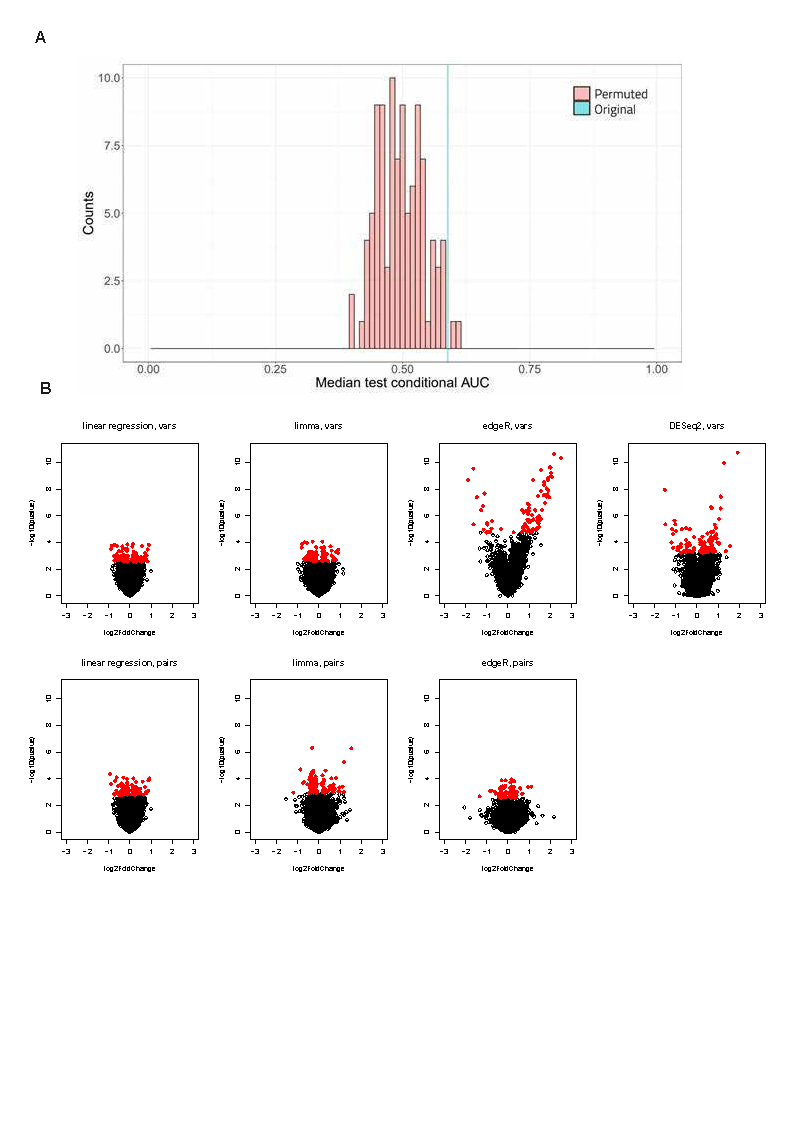

**Extended Data Figure 1: Differential gene expression and model performances in the D-ESMEL discovery set: (A)** The performances of models built on permuted and original data are compared as histogram of medians of test conditional AUCs for the 100 permutations and the median test conditional AUC in the original data (light blue line). **(B)** Volcano plots from seven differential gene expression analysis models comparing metastatic cases and matched controls in the discovery set. Each model was conditioned on matching variables (abbreviated as ‘vars’, top row) and on case-control pairs (abbreviated as ‘pairs’, bottom row). Genes highlighted in red represent the 100 genes with smallest p-values in each model.

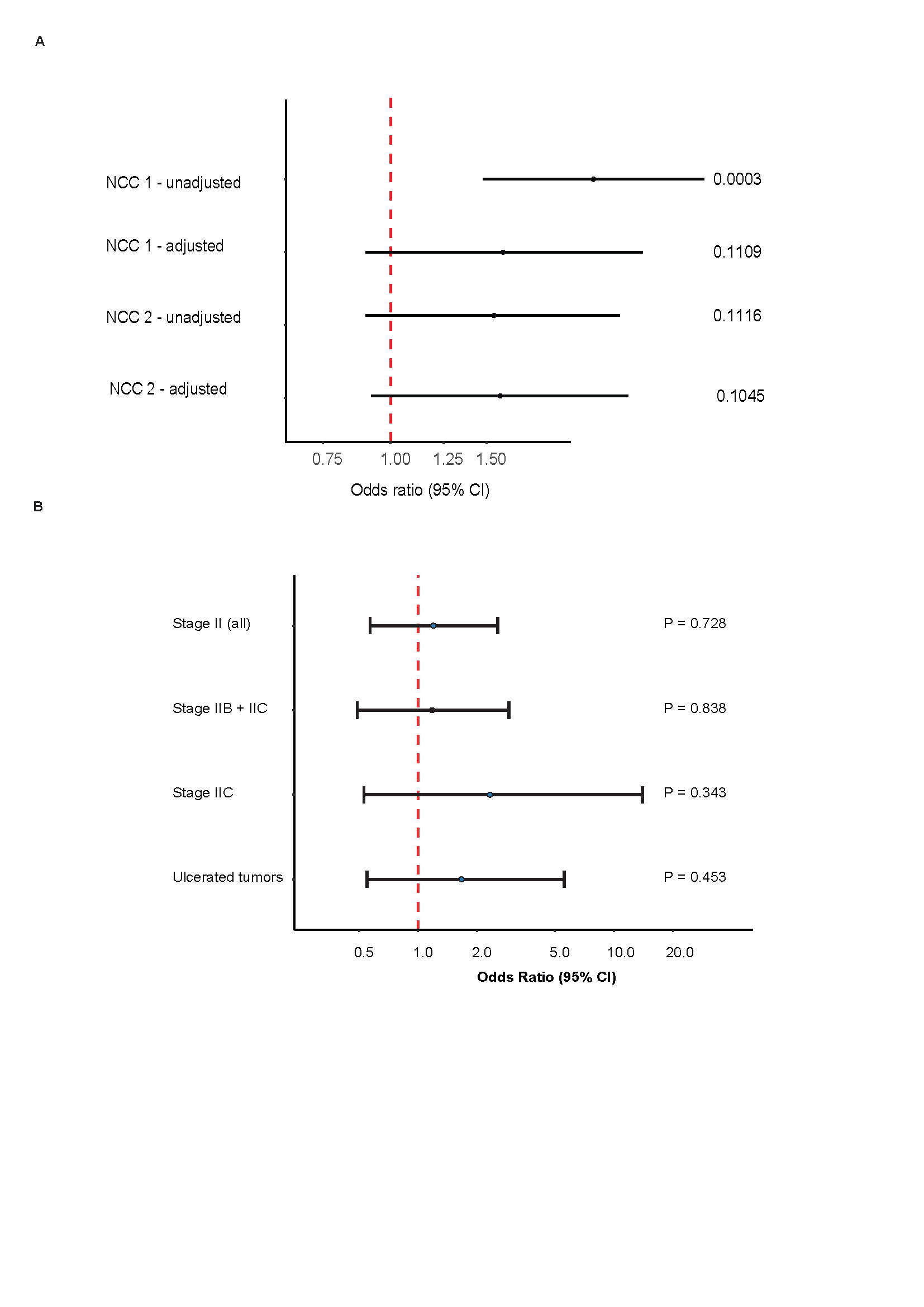

**Extended Data Figure 2: Analyses of clonal loss of p16 with a clinically feasible cut-off value (≥ 20%) and subgroup analyses: (A)** Conditional logistic regression evaluating a ≥ 20% clonal loss of p16 assessed by immunohistochemistry. Univariable analyses with randomly selected controls evaluate the effect of clonal loss of p16 only. Multivariable analyses are adjusted for Breslow thickness, ulceration, age and sex. Univariable analyses with stage-matched controls are controlled for AJCC substage by design. Multivariable analyses are also adjusted for age and sex. **(B)** Subgroup analyses of conditional logistic regression evaluating a ≥ 20% clonal loss of p16 assessed by immunohistochemistry. Subgroup analyses were performed in NCC dataset 2 (stage-matched controls), without adjustment for other variables.

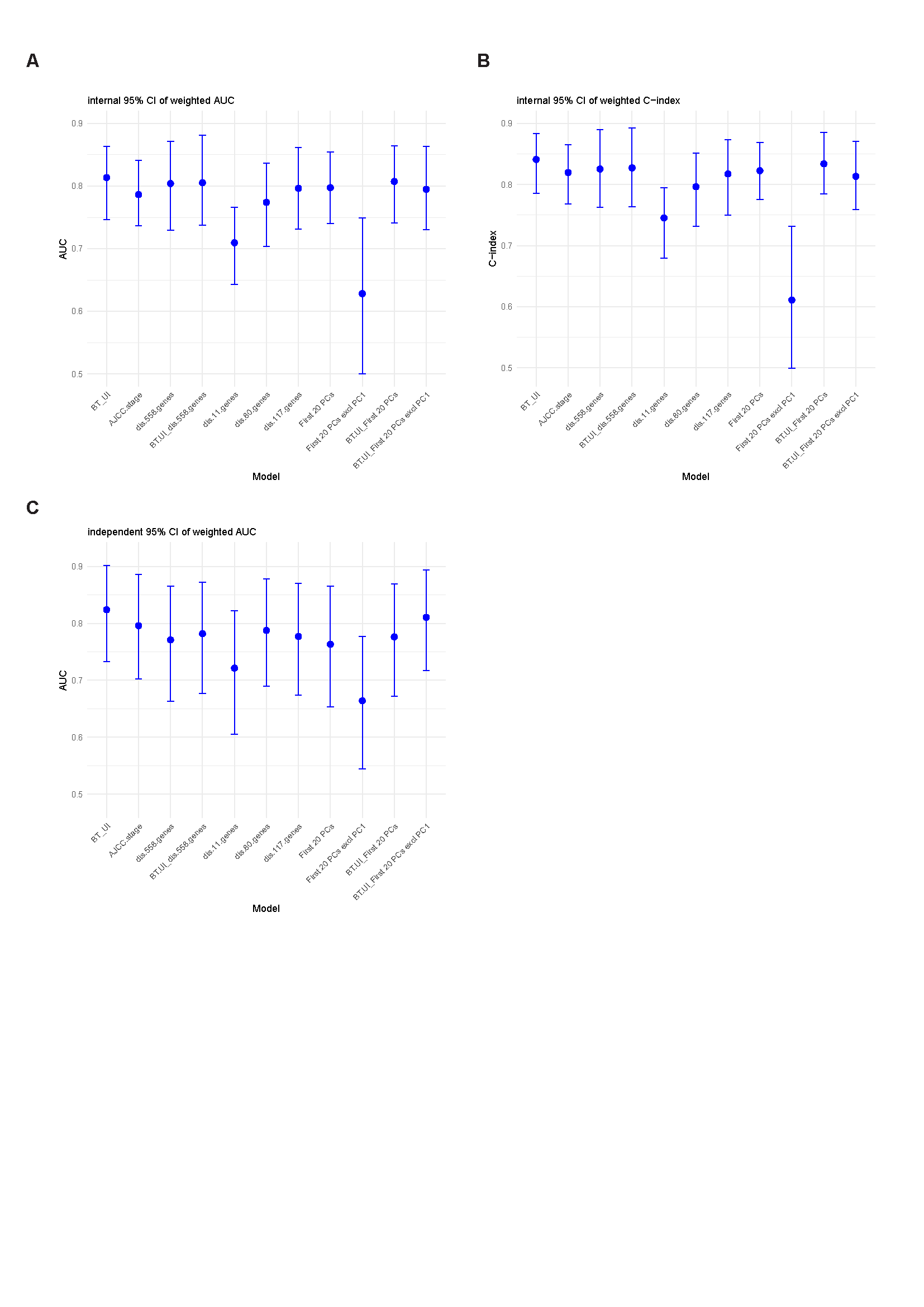

**Extended Data Figure 3: Model performance measurements of gene expression models in the validation cohort: (A)** Weighted area under the curve (AUC) of multiple models developed on the model development subset (i.e. 2/3 of the validation cohort) **(B)** Weighted c-index of multiple models developed on the model development subset **(C)** Weighted AUC of multiple models in the independent validation subset (i.e. 1/3 of the validation cohort, left out during model development).

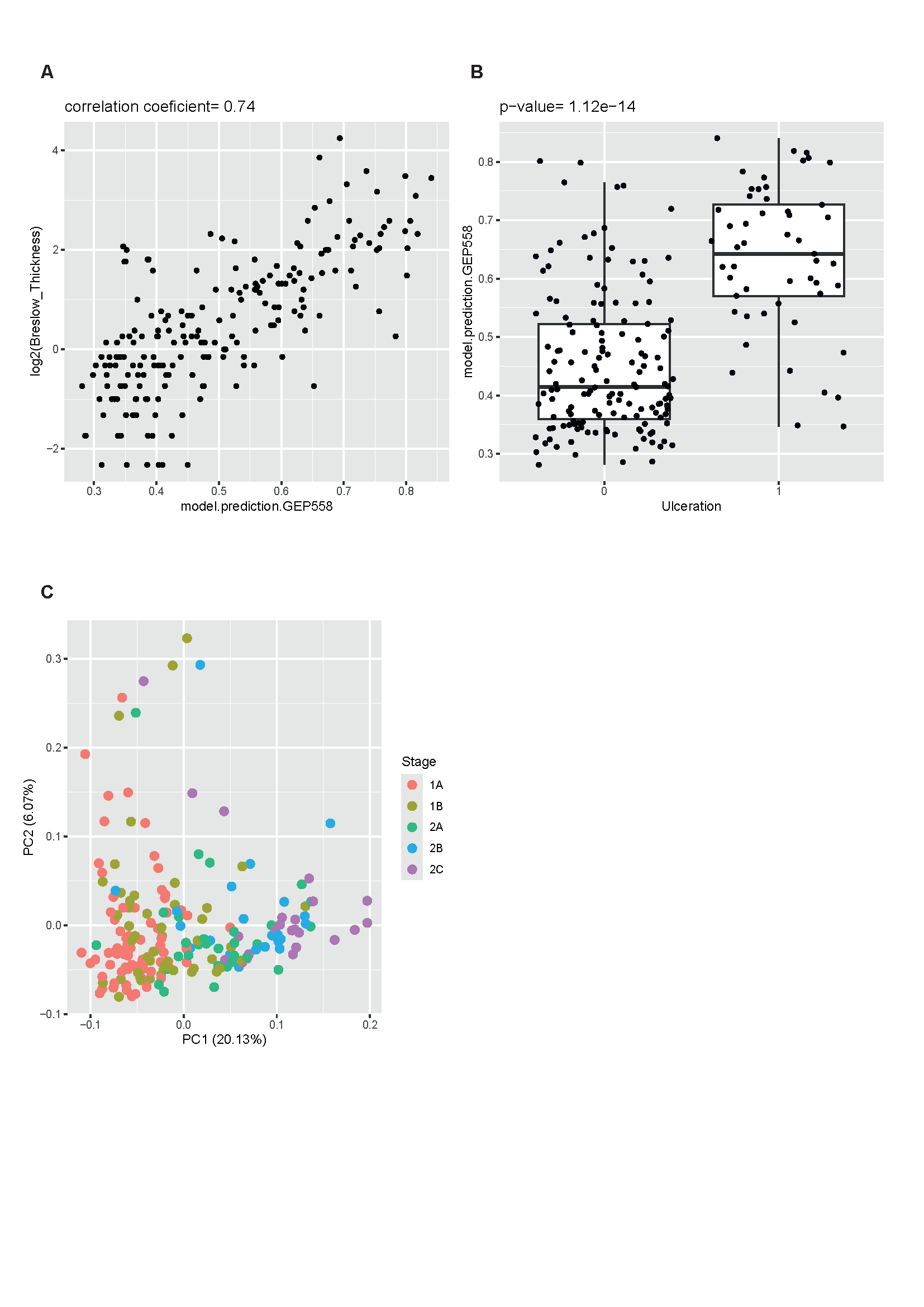

**Extended Data Figure 4: Association between gene expression and staging variables in the D-ESMEL validation cohort: (A)** Association of the 558 discovery genes with Breslow thickness and ulceration in the model development subset of the validation cohort. Correlation between the aggregated prediction score based on the 558 discovery genes and Breslow thickness (log₂-transformed), showing a strong positive correlation (r = 0.74). **(B)** Distribution of prediction scores based on 558 genes by ulceration status, demonstrating significantly higher values in ulcerated compared with non-ulcerated tumors (p = 1.1 × 10⁻¹⁴). **(C)** Principal component analysis (PCA) on the 558 discovery genes in the validation cohort. A scatterplot of PC1 versus PC2 is shown and colored by AJCC stage.

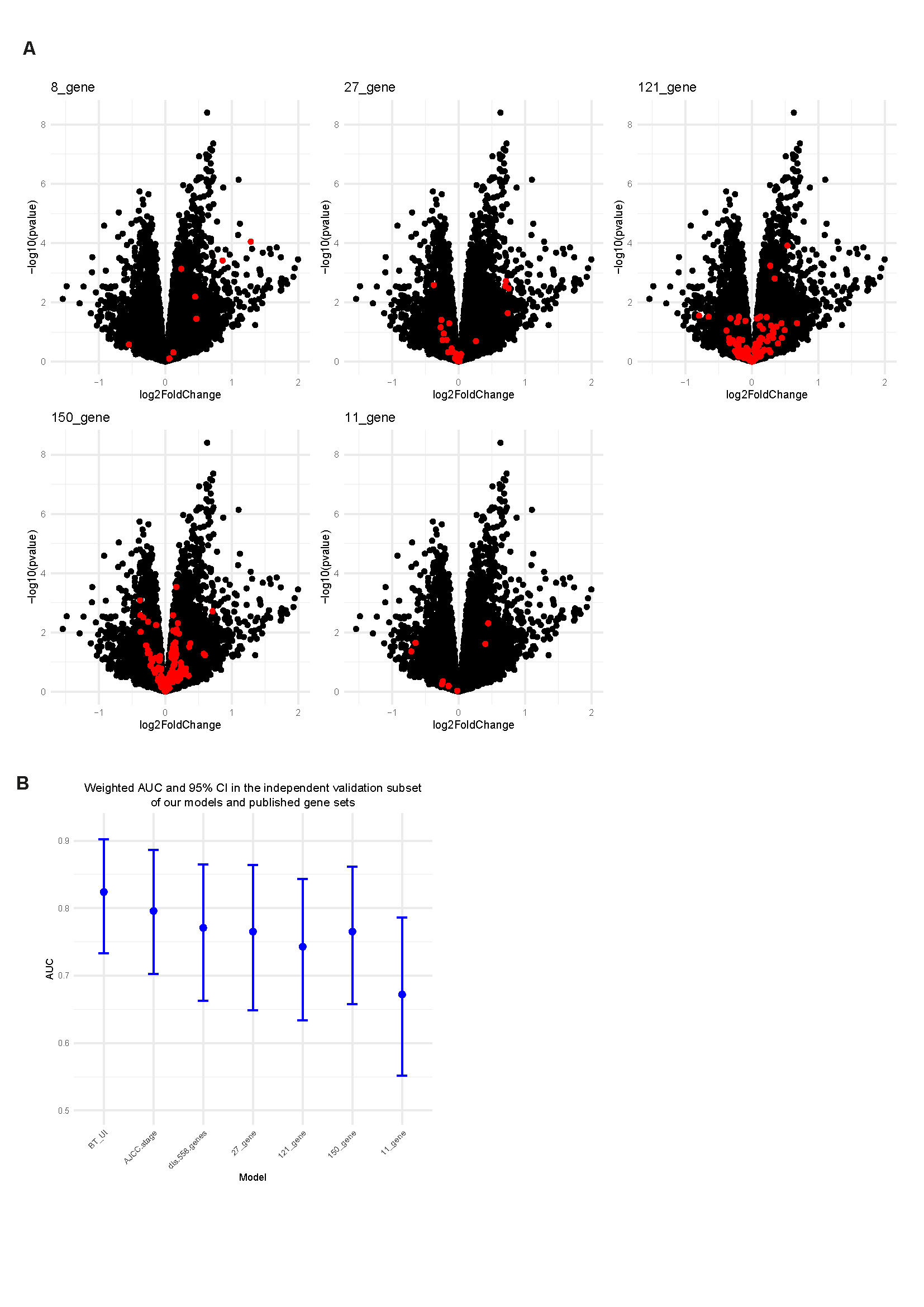

**Extended Data Figure 5: Differential gene expression and model performance measurements of published gene sets in the D-ESMEL validation cohort:** **(A)** Differential expression of genes from published gene sets in the validation cohort. Volcano plots show adjusted p-values and effect sizes for external gene sets. Highlighted in red are overlapping genes from our 558 gene list. Conditioned on age, sex, Breslow thickness, and ulceration. **(B)** Weighted Area Under the Curve (AUC) with 95% confidence intervals based on 1,000 bootstrap iterations (B = 1000) are shown for models based on Breslow thickness and ulceration (BT_Ul), AJCC substage, the 558 discovery genes, and previously published gene sets.
