## Supplementary tables 1-2-4-8-9-10-11-12 for "A Population-Based Multi-Omics Resource for Stage I/II Melanoma Reveals Prognostic Biomarkers but Challenges Their Clinical Utility Beyond Staging"

### Supplementary Table 1: Clinical characteristics of the discovery set of all samples with RNAseq data

|  |  | **Cases** | **Controls** |
| --- | --- | --- | --- |
| N |  | 155 | 147 |
| Age (median (IQR)) |  | 64 (50- 71) | 64 (50- 70) |
| Sex n (%) | Male | 100 (65%) | 91 (62%) |
|  | Female | 55 (35%) | 56 (38%) |
| Breslow thickness (median (IQR)) |  | 2.20 (1.20- 3.40) | 2.10 (1.12- 3.30) |
| Ulceration status | No | 100 (65%) | 95 (65%) |
|  | Yes | 55 (35%) | 52 (35%) |
| stage n (%) | IA | 27 (17%) | 25 (17%) |
|  | IB | 36 (23%) | 40 (27%) |
|  | IIA | 35 (23%) | 29 (20%) |
|  | IIB | 42 (27%) | 36 (24%) |
|  | IIC | 15 (9.7%) | 17 (12%) |
| Histological subtype n (%) | Superficial spreading | 116 (79%) | 111 (79%) |
|  | Lentigo maligna melanoma | 5 (3.4%) | 2 (1.4%) |
|  | Acral melanoma | 4 (2.7%) | 4 (2.8%) |
|  | Nodular | 22 (15%) | 24 (17%) |
|  | NA | 8 | 6 |
| Excision biopsy | Excision | 140 (90%) | 139 (95%) |
|  | biopsy | 15 (9.7%) | 8 (5.4%) |
| Number of histopathologically confirmed metastases per patient n (%) | 1 | 75 |  |
|  | >1 | 5 |  |
| Location of distant metastsis n (%) | lymph node (distant) | 12 |  |
|  | lung | 14 |  |
|  | Gastro-intestinal tract | 12 |  |
|  | Brain | 9 |  |
|  | Bone OR Bone marrow | 6 |  |
|  | Liver | 6 |  |
|  | Skin | 11 |  |
|  | Other | 12 |  |

### Supplementary Table 2. Clinical characteristics of the discovery set of all samples with RNAseq data

| **Characteristic** | **Discovery set (*n* = 356)** | |
| --- | --- | --- |
|  | Cases  (*n* = 178) | Controls  (*n* = 178) |
| **Sex (*n* (%))** |  |  |
| Male | 111 (62) | 111 (62) |
| Female | 67 (38) | 67 (38) |
| **Age, years (median (IQR))** | 67 (50,71) | 63 (52, 71) |
| **Breslow thickness, mm (median (IQR))** | 1.9 (1.0, 3.4) | 1.8 (1.0, 3.3) |
| **Ulcerated (*n* (%))** | 60 (34) | 60 (34) |
| **Stage (*n* (%))** |  |  |
| IA | 33 (19) | 33 (19) |
| IB | 52 (29) | 52 (29) |
| IIA | 30 (17) | 30 (17) |
| IIB | 46 (26) | 46 (26) |
| IIC | 17 (10) | 17 (10) |
| **Body site (*n* (%))** |  |  |
| Face/neck | 11 (6) | 9 (5) |
| Scalp | 20 (11) | 11 (6) |
| Upper extremities | 30 (17) | 41 (23) |
| Trunk | 86 (48) | 77 (43) |
| Lower extremities | 31 (17) | 40 (22) |
| Unknown | 0 (0) | 0 (0) |
| **Morphological subtype (*n* (%))** |  |  |
| Superficial spreading | 118 (66) | 122 (69) |
| Nodular | 40 (22) | 45 (25) |
| Acral lentiginous | 1 (1) | 0 (0) |
| Lentigo maligna melanoma | 2 (1) | 1 (1) |
| Amelanotic melanoma | 1 (1) | 0 (0) |
| Desmoplastic melanoma | 1 (1) | 0 (0) |
| Unspecified | 15 (8) | 10 (6) |
| **Sentinel lymph node biopsy performed (*n*(%))** | 56 (32) | 71 (40) |
| **Time until stage IV recurrence, years (median (IQR))** | 3.5 (1.7, 5.3) | n/a |
| **Time until stage IV recurrence (*n*(%))** |  |  |
| <2 years | 57 (32) | n/a |
| 2-5 years | 67 (38) |  |
| 5-8 years | 42 (24) |  |
| >8 years | 2 (7) |  |
| **Alive survival status at last follow-up (*n*(%))** | 49 (28) | 137 (77) |
| **Follow-up duration until censoring or death, years (median (IQR))** | 5.8 (3.4, 8.9) | 9.7 (7.2, 12.1) |
| **Follow-up duration until censoring or death, years (*n*(%))** |  |  |
| <2 years | 20 (11) | 6 (3) |
| 2-5 years | 52 (29) | 9 (5) |
| 5-8 years | 53 (30) | 42 (24) |
| >8 years | 52 (29) | 121 (68) |
| **Disease progression after initial follow-up period (*n* (%))** |  |  |
| Stage III | n/a | 22 (12) |
| Stage IV | n/a | 18 (10) |

### Supplementary Table 4: Best predictors for metastasis risk in the discovery set of the D-ESMEL study.

| Generalized unconditional regression models adjusted for Breslow thickness, ulceration, age and sex.  Three multivariable models (1 per Level) that were reduced with backward selection | | | | | |
| --- | --- | --- | --- | --- | --- |
| **level** | **Alteration** | **Generalized Linear regression adjusted to the matching variables (139 controls and 138 cases)** | | **conditional logistic regression 9 (91 controls and 91 cases)** | |
| **Copy number variations** | **ARID1B: HD** | 2.2 (1.15-4.40) | 0.02 | 3.27 (1.07-10.01) | 0.04 |
|  | **CDKN2A: HD** | 1,94 (1.17-3.08) | 0.01 | 1.94 (0.99-3.80) | 0.05 |
| **Altered genes** | **ARID1B inactivation** | 2.19 (1.14-4.37) | 0.02 | 3.75 (1.24-11.30) | 0.02 |
|  | **CDKN2A inactivation** | 1.84 (1.14-3) | 0.01 | 2.08 (1.07-4.03) | 0.03 |
| **Pathway level** | **Cell_Cycle** | 1.84 (1.13-3.01) | 0.02 | 2.12 (1.18–3.84) | 0.01 |

### Supplementary Table 8. Clinical characteristics of the validation cohort.

For gene expression analyses the validation cohort was randomly split into two-thirds for model development and one-third for independent model validation.

| **Characteristic** | **Nationwide validation cohort (*n* = 5,551)** | | | | | |
| --- | --- | --- | --- | --- | --- | --- |
|  | **Model development subset (2/3)** | | | **Independent validation subset (1/3)** | | |
|  | Source cohort  (*n* = 3,776) | All cases from source cohort  (*n* = 100) | Randomly selected controls  (*n* = 100) | Source cohort  (*n* = 1,775) | All cases from source cohort  (*n* = 47) | Randomly selected controls  (*n* = 47) |
| **Sex (*n* (%))** |  |  |  |  |  |  |
| Male | 1851 (49) | 52 (52) | 42 (42) | 858 (48) | 27 (57) | 20 (43) |
| Female | 1925 (51) | 48 (48) | 58 (58) | 917 (52) | 20 (43) | 27 (57) |
| **Age, years (median (IQR))** | 63 (51, 73) | 67 (54, 76) | 63 (53, 71) | 62 (51, 73) | 69 (60, 77) | 64 (51, 72) |
| **Breslow thickness, mm (median (IQR))** | 0.8 (0.5, 1.4) | 2.4 (1.3, 4.2) | 0.8 (0.5-1.2) | 0.8 (0.5, 1.4) | 3.8 (1.3, 5.0) | 0.8 (0.5, 1.8) |
| **Ulcerated (*n* (%))** | 355 (9) | 46 (46) | 7 (7) | 158 (9) | 19 (40) | 4 (9) |
| **Stage (*n* (%))** |  |  |  |  |  |  |
| IA | 2347 (62) | 15 (15) | 60 (60) | 1101 (62) | 4 (9) | 25 (53) |
| IB | 767 (20) | 22 (22) | 25 (25) | 370 (21) | 13 (28) | 13 (28) |
| IIA | 322 (9) | 21 (21) | 11 (11) | 140 (8) | 7 (15) | 2 (4) |
| IIB | 212 (6) | 19 (19) | 3 (3) | 112 (6) | 10 (21) | 7 (15) |
| IIC | 128 (3) | 23 (23) | 1 (1) | 52 (3) | 13 (28) | 0 (0) |
| **Body site (*n* (%))** |  |  |  |  |  |  |
| Face/neck | 296 (8) | 7 (7) | 3 (3) | 137 (8) | 6 (13) | 3 (6) |
| Scalp | 194 (5) | 9 (9) | 5 (5) | 114 (6) | 5 (11) | 2 (4) |
| Upper extremities | 872 (23) | 21 (21) | 23 (23) | 393 (22) | 6 (13) | 10 (21) |
| Trunk | 1512 (40) | 38 (38) | 42 (42) | 689 (39) | 17 (36) | 19 (40) |
| Lower extremities | 899 (24) | 25 (25) | 27 (27) | 441 (25) | 13 (28) | 13 (28) |
| Unknown | 3 (0) | 0 (0) | 0 (0) | 1 (0) | 0 (0) | 0 (0) |
| **Morphological subtype (*n* (%))** |  |  |  |  |  |  |
| Superficial spreading | 3042 (81) | 68 (68) | 89 (89) | 1383 (78) | 27 (57) | 36 (77) |
| Nodular | 276 (70 | 24 (24) | 6 (6) | 153 (9) | 16 (34) | 7 (15) |
| Acral lentiginous | 38 (1) | 2 (2) | 0 (0) | 14 (1) | 1 (2) | 0 () |
| Lentigo maligna melanoma | 194 (5) | 2 (2) | 1 (1) | 107 (6) | 2 (4) | 1 (2) |
| Amelanotic melanoma | 4 (0) | 0 (0) | 1 (1) | 2 (0) | 0 (0) | 1 (2) |
| Desmoplastic melanoma | 24 (1) | 0 (0) | 1 (1) | 10 (1) | 0 (0) | 0 (0) |
| Unspecified | 198 (5) | 4 (4) | 2 (2) | 106 (6) | 1 (2) | 2 (4) |
| **Sentinel lymph node biopsy performed (*n*(%))** | 1185 (31) | 47 (47) | 30 (30) | 598 (34) | 21 (45) | 16 (34) |
| **Time until stage IV recurrence, years (median (IQR))** | n/a | 2.0 (1.1, 3.4) | n/a | n/a | 1.6 (1.0, 3.2) | n/a |
| **Time until stage IV recurrence (*n*(%))** |  |  |  |  |  |  |
| <2 years | n/a | 51 (51) | n/a | n/a | 29 (62) | n/a |
| 2-5 years |  | 46 (46) |  |  | 18 (38) |  |
| 5-8 years |  | 3 (3) |  |  | 0 (0) |  |
| >8 years |  | 0 (0) |  |  | 0 (0) |  |
| **Alive survival status at last follow-up (*n*(%))** | 3446 (91) | 30 (30) | 93 (93) | 1628 (92) | 12 (26) | 43 (92) |
| **Follow-up duration until censoring or death, years (median (IQR))** | 3.4 (2.2, 4.7) | 3.7 (2.1, 5.5) | 6.1 (5.0, 7.2) | 3.4 (2.2, 4.6) | 3.4 (1.8, 5.1) | 6.1 (4.5, 7.3) |
| **Follow-up duration until censoring or death, years (*n*(%))** |  |  |  |  |  |  |
| <2 years | 763 (20) | 24 (24) | 1 (1) | 392 (22) | 15 (32) | 1 (2) |
| 2-5 years | 2251 (20) | 44 (44) | 24 (24) | 1052 (59) | 20 (43) | 14 (30) |
| 5-8 years | 762 (20) | 32 (32) | 75 (75) | 331 (19) | 12 (26) | 31 (66) |
| >8 years | 0 (0) | 0 (0) | 0 (0) | 0 (0) | 0 (0) | 1 (2) |
| **Disease progression after initial follow-up period (*n* (%))** |  |  |  |  |  |  |
| Stage III | n/a | n/a | 4 (4) | n/a | n/a | 1 (1) |
| Stage IV | n/a | n/a | 1 (1) | n/a | n/a | 0 (0) |

### Supplementary Table 9. Performance of prognostic models during model development and independent validation.

Weighted AUC and C-index are shown for each model, using 0.632+ correction in the model development subset.

| **Model** | **Features** | **Model development subset** | | **Independent validation subset** | |
| --- | --- | --- | --- | --- | --- |
|  |  | **Weighted AUC** | **Weighted C-index** | **Weighted AUC** | **Weighted C-index** |
| Clinicopathological | Breslow thickness*, ulceration | 0.82 (95% CI 0.78-0.86) | 0.85 (95% CI 0.72-0.88) | 0.82 (95% CI 0.73-0.90) | 0.84 (95% CI 0.76-0.91) |
| AJCC stage | Stage IA, IB, IIA, IIB, IIC | 0.79 (95% CI 0.75-0.82) | 0.83 (95% CI 0.79-0.86) | 0.80 (95% CI 0.70-0.89) | 0.81 (95% CI 0.72-0.89) |
| Matching variables | Age*, sex, Breslow thickness*, and ulceration | 0.79 (95% CI 0.74-0.83) | 0.83 (95% CI 0.78-0.86) | 0.81 (95% CI 0.72-0.89) | 0.83 (95% CI 0.75-0.90) |
| GEP | 558 discovery genes all tumors | 0.79 (95% CI 0.74-0.88) | 0.82 (95% CI 0.77-0.89) | 0.77 (95% CI 0.66-0.86) | 0.79 (95% CI 0.69-0.88) |
|  | 558 discovery genes in stage I | 0.72 (95% CI 0.54-0.81) | 0.72 (95% CI 0.54-0.82) | 0.69 (95% CI 0.52-0.84) | 0.64 (95% CI 0.46-0.80) |
|  | 558 discovery genes in stage II | 0.47 (95% CI 0.20-0.75) | 0.54 (95% CI 0.27-0.77) | 0.46 (95% CI 0.25-0.70) | 0.48 (95% CI 0.26-0.71) |
| Clinicopathological + GEP | Breslow thickness*, ulceration, 558 genes from discovery | 0.80 (95% CI 0.74-0.88) | 0.83 (95% CI 0.77-0.89) | 0.79 (95% CI 0.68-0.87) | 0.81 (95% CI 0.71-0.89) |
| High selection frequency GEP | 11 genes | 0.71 (95% CI 0.65-0.77) | 0.75 (95% CI 0.70-0.81) | 0.72 (95% CI 0.61-0.82) | 0.75 (95% CI 0.64-0.84) |
|  | 80 genes | 0.79 (95% CI 0.71-0.85) | 0.82 (95% CI 0.72-0.85) | 0.79 (95% CI 0.69-0.88) | 0.80 (95% CI 0.69-0.89) |
|  | 117 genes | 0.80 (95% CI 0.74-0.86) | 0.82 (95% CI 0.76-0.87) | 0.78 (95% CI 0.67-0.87) | 0.80 (95% CI 0.70-0.88) |
| Principal Component Analyses | First 20 PCs  First 20 PCs excluding PC1  Clinicopathological+ First 20 PCs  Clinicopathological+ First 20 PCs excluding PC1 | 0.80 (95% CI 0.74-0.85)  0.66 (95% CI 0.50-0.74)  0.81 (95% CI 0.75-0.87)  0.81 (95% CI 0.73-0.85) | 0.83 (95% CI 0.78-0.87)  0.63 (95% CI 0.50-0.72)  0.84 (95% CI 0.70-0.88)  0.83 (95% CI 0.76-0.86) | 0.77 (95% CI 0.65-0.87)  0.62 (95% CI 0.54-0.78)  0.78 (95% CI 0.67-0.87)  0.76 (95% CI 0.72-0.89) | 0.78 (95% CI 0.68-0.87)  0.62 (95% CI 0.55-0.78)  0.79 (95% CI 0.68-0.87)  0.77 (95% CI 0.73-0.89) |
| GEP based on model development subset of the validation cohort | 50 genes | 0.80 (95% CI 0.73-0.86) | 0.81 (95% CI 0.74-0.86) | 0.79 (95% CI 0.68-0.88) | 0.81 (95% CI 0.71-0.89) |
|  | 100 genes | 0.80 (95% CI 0.73-0.87) | 0.81 (95% CI 0.75-0.87) | 0.78 (95% CI 0.68-0.88) | 0.80 (95% CI 0.71-0.88) |
|  | 200 genes | 0.81 (95%CI 0.74-0.87) | 0.81 (95% CI 0.75-0.87) | 0.78 (95% CI 0.67-0.88) | 0.80 (95% CI 0.70-0.88) |
|  | 500 genes | 0.82 (95% CI 0.74-0.87) | 0.82 (95% CI 0.75-0.87) | 0.77 (95% CI 0.66-0.87) | 0.80 (95% CI 0.70-0.89) |
|  | 1000 genes | 0.82 (95% CI 0.73-0.87) | 0.83 (95% CI 0.74-0.88) | 0.77 (95% CI 0.76-0.88) | 0.80 (95% CI 0.70-0.86) |

*Variable was included as a continuous variable. Abbreviations: AJCC = American Joint Committee of Cancer, GEP = gene expression profile.

### Supplementary Table 10 Pathway Analyses

| **Pathways** | **Odds Ratio (95% CI)** | **P value** |
| --- | --- | --- |
| WNT | 0.645 (0.221–1.886) | 0.423 |
| RTK_RAS | 0.574 (0.273–1.203) | 0.141 |
| PI3K | 1.277 (0.577–2.824) | 0.546 |
| TP53_DNA_damage | 1.446 (0.725–2.887) | 0.295 |
| Cell_Cycle | 2.124 (1.176–3.836) | 0.012 |
| MYC | 0.389 (0.074–2.048) | 0.265 |
| TGF_beta | 1.138 (0.420–3.081) | 0.800 |

### Supplementary Table 11: Definition of CNV based on RPC and SNPs

| **Alteration Type** | **Mean RPC Range** | **SNP Condition** |
| --- | --- | --- |
| Homozygous Deletion (HD) | < -50 | All SNPs in the gene are imbalanced |
| Loss of Heterozygosity (LOH) | > -50 and < -10 | More than 2 SNPs are imbalanced |
| Gain | > 10 and < 100 | No specific SNP condition required |

### Supplementary Table 12: Criteria used to determine clonal relatedness between primary tumors and metastases based on targeted sequencing

| **Genomic findings** | **Interpretation** |
| --- | --- |
| ≥3 identical CNVs between samples | Clonally related |
| Identical non-hotspot mutations + ≥1 identical CNV | Clonally related |
| ≥1–2 identical CNVs between samples | Likely clonally related |
| Identical non-hotspot mutations shared between samples | Likely clonally related |
| Absence of hotspot mutations in BRAF or NRAS and/or absence of other mutations in metastasis (while detected in the primary tumor) | Not clonally related |
| Only BRAF V600 or other hotspot mutation, or presence of TERT promoter mutation | No conclusion |
| Absence of detectable mutations or CNVs in the primary tumor (e.g., low tumor cell percentage) | No conclusion |
| Poor sequencing quality (no coverage of hotspot regions and/or unreliable CNV calls) | No conclusion |
