## Supplementary figures for "A Population-Based Multi-Omics Resource for Stage I/II Melanoma Reveals Prognostic Biomarkers but Challenges Their Clinical Utility Beyond Staging"

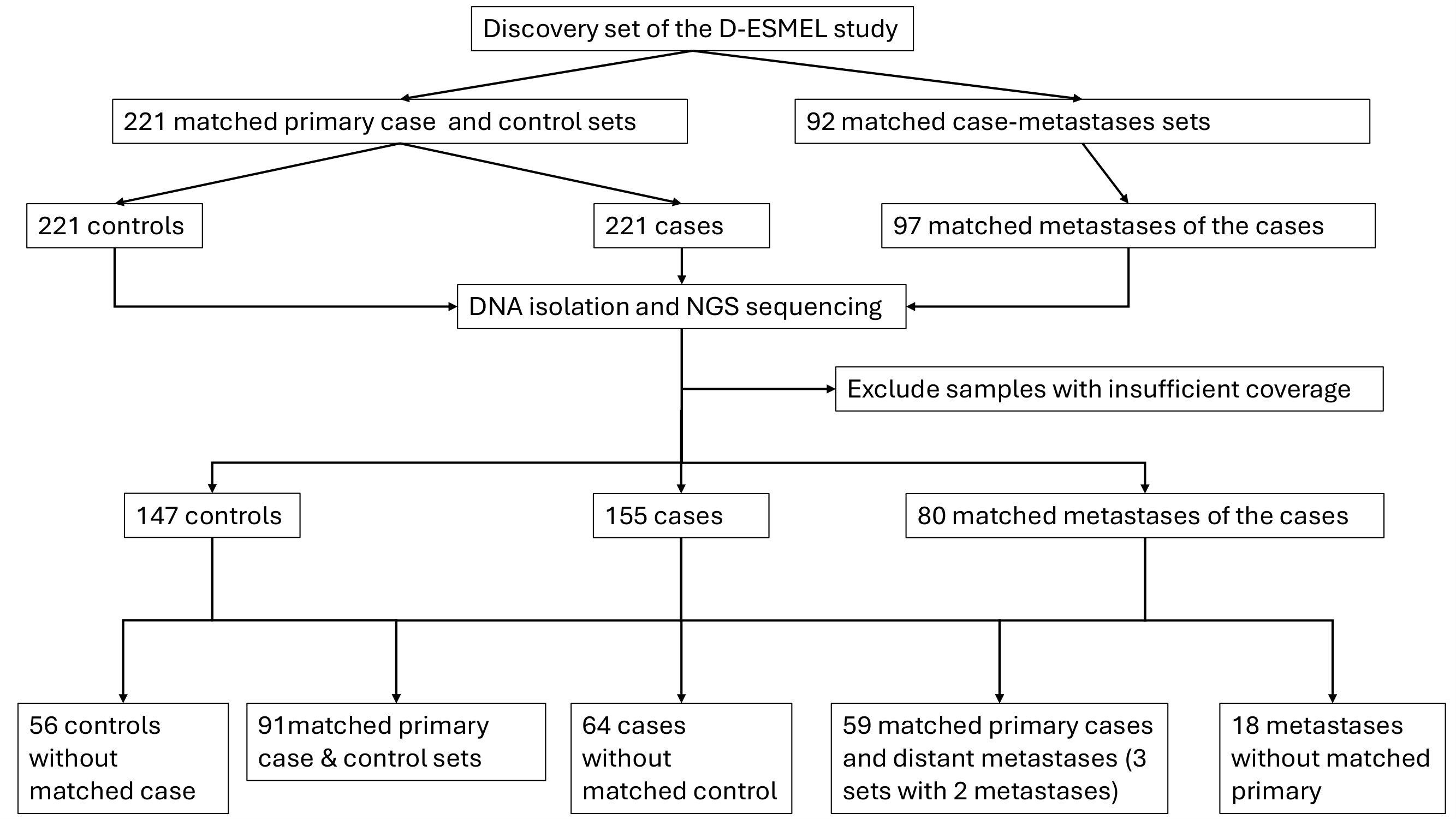

### **Figure S1. Flow chart of sample collection for genomic analysis in the D-ESMEL study.** The discovery cohort comprised 221 matched primary melanoma case-control sets and 92 matched case-metastasis sets, including 97 matched metastatic samples. Following DNA isolation and next-generation sequencing (NGS), samples with insufficient sequencing coverage were excluded, resulting in 147 controls, 155 primary melanoma cases, and 80 matched metastatic samples. The resulting dataset included 91 matched primary case-control sets, 56 controls without a matched case, 64 cases without a matched control, 59 matched primary case–metastasis pairs (including three sets with two metastatic samples), and 18 metastatic samples without a matched primary tumour.

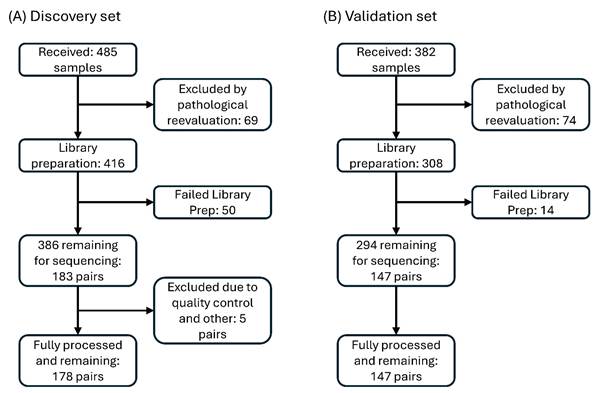

### **Supplementary Figure 2. Flowchart of sample processing and sample exclusion for RNA sequencing.** (A) Discovery set: Of the 485 samples received, 69 were excluded following pathological reevaluation. Of the remaining 416 samples, 50 failed library preparation, leaving 386 samples (183 pairs) for sequencing. A further five pairs were excluded due to quality control (QC) failure and other reasons, resulting in 178 fully processed pairs. (B) Validation set: Of the 382 samples received, 74 were excluded following pathological reevaluation. Of the remaining 308 samples, 14 failed library preparation, leaving 294 samples (147 pairs) for sequencing. All 147 pairs were fully processed and retained for downstream analysis. The validation set includes the model development and independent validation subsets.

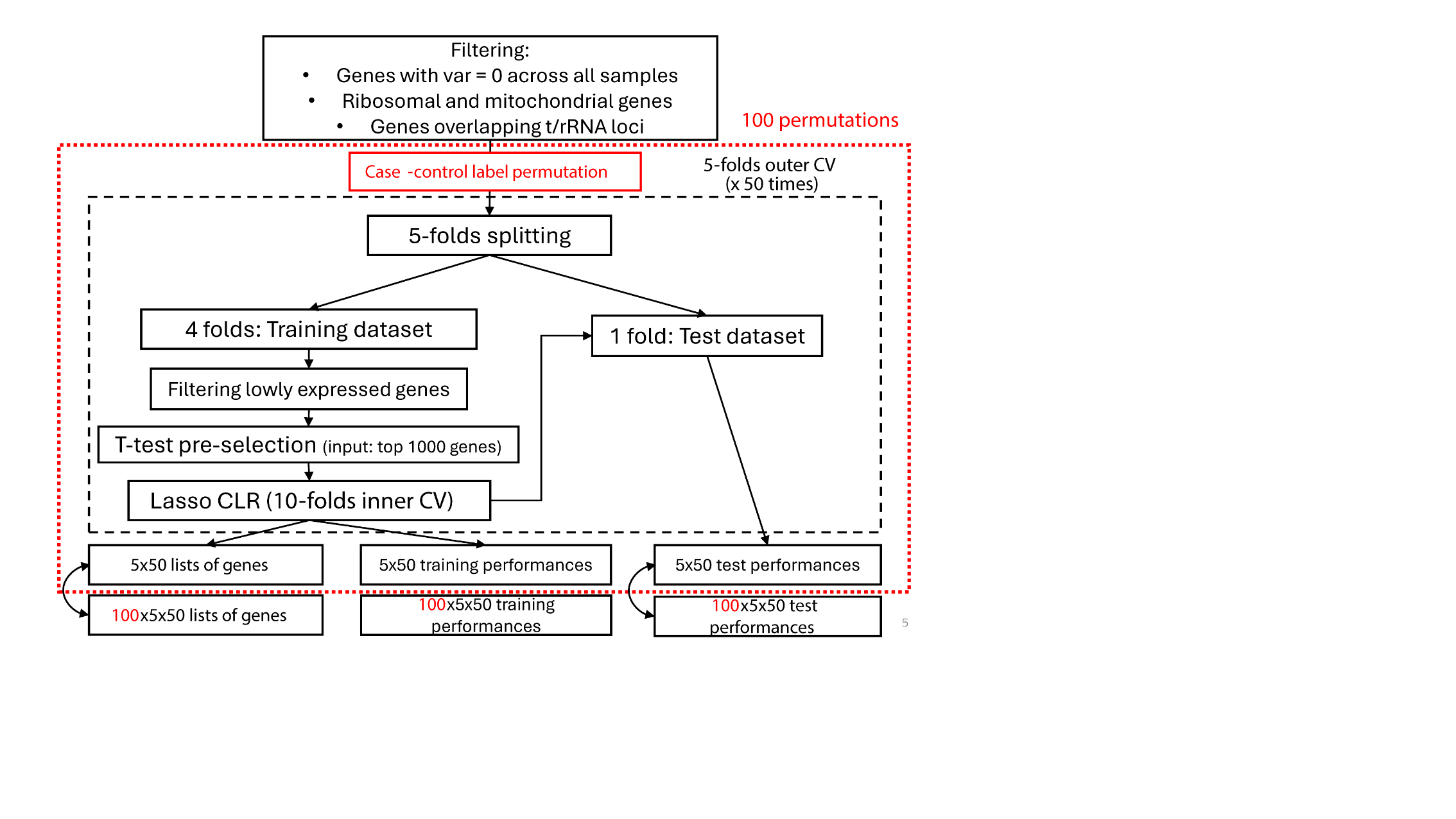

### **Supplementary Figure 3 Schematic of the double-loop cross-validation (DLCV) procedure used for gene selection and model evaluation.**

After filtering, the dataset (n = 356) was split into 5 outer folds, repeated 50 times. Within each training set, lowly expressed genes were removed, and paired t-tests were used for pre-selection. LASSO-penalized conditional logistic regression with 10-fold inner cross-validation was applied. Model performance was assessed on the outer test folds. To evaluate robustness of selected features, case-control labels were permuted 100 times.
